# Wastewater Surveillance as an Event Detection System: Outbreak and Peak Detection of SARS-CoV-2 Across 281 U.S. Counties

**DOI:** 10.64898/2026.05.14.26353186

**Authors:** Nicholas B Link, Raul Garrido, Anjalika Nande, Mauricio Santillana

**Affiliations:** Network Science Institute, Northeastern University; Physics Department, Northeastern University; Nuffield Department of Primary Care Health Sciences, University of Oxford; Institute for Computational Medicine, Johns Hopkins University

**Keywords:** wastewater-based surveillance, COVID-19, outbreak detection, peak detection, infectious disease surveillance, Bayesian modeling

## Abstract

Wastewater-based surveillance (WBS) is increasingly used to monitor infectious disease dynamics, yet most evaluations focus on correlation or forecasting—neither of which directly assesses whether wastewater signals can identify the epidemiological events most relevant to public health decision-making. We argue that outbreak onset and epidemic peak detection are the operationally critical use cases of WBS, requiring a fundamentally different evaluation framework. We introduce a classification-based framework that treats WBS as an event-detection problem, defining outbreaks and peaks as discrete events, establishing detection intervals to account for timing uncertainty, and incorporating censoring and data completeness criteria for valid comparisons against imperfect clinical reference outcomes. Within this framework, we apply a Bayesian exponential growth model for outbreak detection - benchmarked against a standard reproductive number (Rt)-based method - and a rule-based algorithm for peak detection, evaluating performance via sensitivity and positive predictive value (PPV). Applied to county-level SARS-CoV-2 wastewater data from 281 U.S. counties (Biobot, 2021–2024), the exponential growth approach substantially outperforms the Rt-based baseline: sensitivity 0.82 and PPV 0.64 versus sensitivity 0.58 and PPV 0.19 for the best-performing Rt variant. Peak detection achieves sensitivity 0.84 and PPV 0.70 at the county level. Both peak and outbreak detection achieve strong and consistent performance against hospitalizations and deaths at the state level. Spatial aggregation yields a statistically significant improvement in peak detection PPV against a curated reference standard (*p <* 0.001), while outbreak detection improvements under aggregation are directionally consistent but not statistically significant. Wastewater leads case-defined outbreaks by 4–6 days but minimally leads epidemic peaks, consistent with wastewater approximating prevalence rather than incidence. These findings demonstrate that wastewater signals can reliably detect outbreak onset and epidemic peaks across spatial scales and clinical outcomes, and that the choice of detection method matters substantially in practice. The classification framework developed here provides a reusable and principled tool for evaluating any surveillance signal as an event-detection system, with direct relevance to how WBS is actually used in public health decision-making.

**Highlights:**

- We evaluate wastewater surveillance as an event-detection system for outbreak onset and epidemic peak timing.
- We introduce a classification-based framework that accounts for timing uncertainty, censoring, and data completeness.
- Wastewater signals detect case-defined outbreaks and peaks with strong sensitivity and positive predictive value across spatial scales.
- Peak and outbreak detection show modest gains under aggregation, particularly for noisier outcomes such as deaths.
- The proposed framework provides a reusable approach for evaluating surveillance signals against epidemiologically meaningful events.

## 1 Introduction

Wastewater-based surveillance (WBS)—which monitors diseases through viral shedding in wastewater—has become a widely adopted tool for detecting infectious disease outbreaks and tracking epidemic peaks. The COVID-19 pandemic underscored the need for flexible, scalable monitoring systems that can track both emerging and established pathogens, especially as climate change and global interconnectedness reshape the infectious disease landscape [1]. WBS provides a cost-effective alternative to traditional clinical surveillance, which is often delayed, resource-intensive, and subject to biases introduced by testing availability, health-seeking behavior, and changes in diagnostic tools. WBS enables anonymous and indirect surveillance without requiring individual testing. Although relatively new, WBS scaled rapidly during the COVID-19 pandemic and has demonstrated utility across diverse geographic settings and pathogen types [2].

Public health surveillance systems are often used to identify actionable events—such as the onset of an outbreak or the timing of an epidemic peak—that can inform interventions, guide resource allocation, and support risk communication. Wastewater surveillance has been increasingly used in this way, providing early warning signals of rising transmission that can precede clinical indicators and enable an earlier response [3]. However, its capacity to accurately detect outbreaks and peaks, and serve as an early warning system—particularly across different spatial scales—remains understudied.

During the COVID-19 pandemic, numerous studies found strong correlations between SARS-CoV-2 wastewater viral concentrations and clinical outcomes such as new cases, hospitalizations, and deaths [4, 5]. These findings suggest that wastewater data can serve as a proxy for underlying incidence (the true, unobserved number of new infections) partially reflected in clinical data. Many studies have observed that correlations improve when a time lag is introduced, with wastewater measurements preceding clinical outcomes, highlighting the potential of WBS as an early indicator of outbreaks [4]. However, correlation analyses have key limitations: they are sensitive to long-term shifts in the time series (e.g. the rise of at-home testing in 2022 [6]) and they do not directly evaluate whether wastewater can detect outbreaks or peaks in a timely and reliable manner. Outbreak and peak detection are a distinct and essential use case of WBS, with implications for rapid response and resource allocation.

WBS is already used in practice as an outbreak monitoring tool. For example, the Boston Public Health Commission has implemented neighborhood-level wastewater sampling to detect local shifts in transmission [7]. Some studies have shown that wastewater trends anticipate outbreaks in case data [8] and peaks in hospitalization data [9], likely due to wastewater’s lead time over clinical outcomes. In addition to identifying outbreak onset, detecting epidemic peaks is also operationally relevant: peak timing can inform when transmission has reached its maximum, support hospital capacity planning, and guide the scaling up or down of public health interventions. Wastewater signals, which reflect upstream transmission dynamics, may provide a timely indicator of peak timing even when clinical data are delayed or incomplete.

Wastewater has been widely used to forecast clinical outcomes using statistical and machine learning models [4, 10]. Forecasting is an important component of epidemic analysis, but it is also a challenging task that often requires strong assumptions and can perform inconsistently in practice, particularly in the presence of noisy or rapidly changing data; state-of-the-art forecasting models have repeatedly struggled to anticipate sharp increases in disease activity in real-time operational settings [11]. In contrast, detecting epidemiological events such as outbreak onset or peak timing reframes the problem into identifying meaningful changes in observed signals. This can be a more attainable objective while remaining closely aligned with public health decision-making. Rather than replacing forecasting, event detection provides a complementary perspective that focuses on identifying when conditions warrant attention or action.

Despite the practical importance of outbreak and peak detection, there has been limited work formalizing how to evaluate the ability of wastewater signals to detect such events. Existing studies have characterized technical sources of uncertainty in wastewater signal quality [12]. However, none have systematically developed and evaluated a classification-based framework for outbreak and peak detection using wastewater at population scale —-one that quantifies true positive rates, false alarm rates, and missed detections against empirically defined clinical reference outcomes across diverse settings. For WBS to credibly support public health decision-making, these questions need explicit answers: How many true outbreaks will the system detect? How reliably can wastewater track epidemic peaks? How often will it raise false alarms? The operational questions that public health practitioners face — is an outbreak beginning? has transmission peaked? — are fundamentally detection problems, not trajectory prediction problems. Yet evaluation frameworks in forecasting and predictive modeling are oriented toward predictive accuracy rather than event detection. In this study, we adapt the evaluation perspective from forecasting to the problem of outbreak and peak detection, developing a classification-based framework more directly aligned with the practical use of surveillance systems. Conceptually, this approach is analogous to evaluating a diagnostic test: the goal is to assess how well an observed signal identifies an underlying condition, while recognizing that the reference standard may itself be imperfect. Spatial scale adds an important layer of variability in WBS effectiveness. Surveillance systems operate at different levels — from neighborhood catchment areas to counties, states, and national systems. Aggregating data across larger areas can smooth noise and enhance signal stability, but may also obscure heterogeneity in local transmission. While some studies have examined spatial relationships and clustering in WBS signals [16], few have examined how spatial aggregation affects the accuracy or timeliness of event detection. This study makes three contributions. First, we introduce a classification-based framework for evaluating wastewater surveillance as an event-detection system — defining outbreaks and peaks as discrete classifiable events, specifying detection intervals to account for timing uncertainty, and incorporating censoring and data completeness criteria to ensure valid comparisons against imperfect clinical reference outcomes. This framework is the primary methodological contribution of the paper, and it is designed to generalize beyond COVID-19 and beyond wastewater to any surveillance signal evaluated as a detector of epidemiologically meaningful events. Second, we apply this framework empirically, benchmarking a Bayesian exponential growth model against a standard Rt-based baseline for outbreak detection, and evaluating a rule-based peak detection algorithm, demonstrating that simpler and more parsimonious methods can outperform more complex ones when matched to the data structure and the detection objective. Third, we provide a systematic empirical assessment of detection performance across methods, spatial scales, and clinical outcomes — cases, hospitalizations, and deaths — highlighting when wastewater acts as a leading indicator and when it does not, and what that distinction implies for surveillance system design.

## 2 Data

### 2.1 County-level wastewater and case data

We analyze weekly averaged wastewater SARS-CoV-2 data from Biobot, a company specializing in wastewater surveillance. Data were generated through qPCR analysis of wastewater samples that were collected using an autosampler. The sampler collected data once per hour over 24 hour periods. qPCR methodology followed industry standards and consequently evolved over the course of the sampling period. Biobot accounted for biases inherent to methodological variation. Wastewater flow can vary depending on a variety of factors, including rain events, thus accounting for variation in the human contribution across sampling periods is critical. Biobot followed a standard approach to normalize SARS-CoV-2 data using concentrations of the pepper mild mottle virus, which is a ubiquitous virus ingested by humans that eat pepper-containing products [13].

The dataset contains weekly-aggregated data from January 6, 2021, to June 26, 2024, and includes samples from 304 U.S. counties. The specific date range of available data varies by county. To ensure data quality, we restrict our analysis to counties within U.S. states that reported at least 20 weeks of wastewater data prior to May 10, 2023 (shortly after the last reporting of the case data). This filtering step yields 281 counties for analysis.

To handle missing values, we apply linear interpolation for gaps of three or fewer consecutive missing weeks. While a few locations reported data bi-weekly—leading to more frequent interpolation—most counties had only minor gaps. Overall, just 5.5% of all data points were imputed.

We use daily reported COVID-19 case counts from Johns Hopkins University [14]. We correct any days with negative reported cases by reallocating prior counts forward in time. To align with the wastewater data, we aggregate the daily cases into weekly counts. Occasionally, some weeks have no reported cases surrounded by weeks with high case counts. We interpret these as reporting errors and smooth the time series by redistributing neighboring cases across the missing weeks. In contrast, weeks with zero reported cases surrounded by weeks with zero or low case counts are interpreted as accurately reflecting low incidence.

### 2.2 Aggregated and state-level data

In addition to **county-level** data for wastewater and cases, we generate **county-aggregated** and **state-level** data. County aggregated data include only counties with wastewater sampling, whereas state-level data include all counties within the state. To construct county-aggregated wastewater data, we compute a population-weighted average of county-level wastewater measurements at each time point, weighting by the county population. Because the county-level wastewater data are normalized to a common scale across locations, population weighting reflects the relative contribution of each county incidence to the state-level incidence. The corresponding county-aggregated case data are filtered to include only counties with available wastewater data and then aggregated within each state. To generate the state-level data, cases from all counties in a state—even ones without any wastewater sampling— are aggregated. Because state-level wastewater data are not available, we use county-aggregated data as a proxy, reflecting a common use case in which incomplete sampling coverage is still used to monitor state-level trends.

### 2.3 Hospitalizations and deaths

In addition to wastewater and case data, we use weekly hospitalization and death data. Death data are sourced from Johns Hopkins University [14] and aggregated in the same manner as cases to generate county-aggregated and state-level data. Hospitalization data are taken directly from Our World in Data [15], where they are available only at the state-level.

## 3 Methods

This study has a 3-stage design: (1) applying event-detection approaches to wastewater and clinical outcome data, (2) classifying wastewater outbreak and peak detections against outcome-defined events using the proposed evaluation framework, and (3) analyzing how surveillance performance changes under spatial aggregation. The approaches are chosen for interpretability and practical applicability, while still capturing key strategies for event detection in noisy epidemiological data. The onset detection approach uses a Bayesian exponential growth model combined with posterior decision criteria to identify outbreak onset, while the peak detection approach offers a simple and effective rule-based method for identifying epidemic peaks. We compare the onset detection approach against a reproductive number (*R*_*t*_)-based baseline; full details of this comparison are provided in Appendix Section S2. The primary methodological contribution of the paper is the classification-based evaluation framework used to assess event detection performance.

### 3.1 Onset Detection: Exponential Growth Model and Decision Criteria

In the early stages of an outbreak, infectious diseases often exhibit exponential growth—indeed, this is the mechanism in susceptible-infected-recovered (SIR) models. Exponential growth models have already been applied to wastewater data for prediction [16] and have been used for outbreak detection with other data streams [17, 18].

For each location *i* and time *t*, we fit the following exponential growth model:

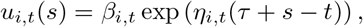

where:

- *u*_*i,t*_(*s*) is the expected value of the outcome (e.g., wastewater RNA, cases, or deaths) at time *s* for the model fit at time *t* and location *i*,
- *β*_*i,t*_ is the intercept of the exponential growth model at time *t* for location *i*, representing *u*_*i,t*_(*t* − *τ*), the expected outcome at the start of the time window,
- *η*_*i,t*_ is the growth rate per unit of time at time *t* for location *i*.

The model is fit independently for each time window *s* ∈ [*t − τ, t*], where *τ* is the width of the time window, across time points *t* ∈ [*τ* + 1, *T*_*i*_], where *T*_*i*_ is the maximum time at location *i*. The model is also fit independently across locations.

Although the assumption of spatio-temporal independence is likely violated in the data, we adopt this framework to focus on detecting spatially and temporally localized outbreaks without introducing global spatio-temporal smoothing. The outcome distribution for each location *i* is assumed to follow a Negative Binomial distribution, which is appropriate for non-negative count data that may exhibit overdispersion:

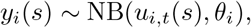

where:

- *y*_*i*_(*s*) is the observed outcome at time *s* for location *i*,
- *u*_*i,t*_(*s*) is the expected value of the Negative Binomial distribution at time *s* for the model fit at time *t* for location *i*,
- *θ*_*i*_ is the dispersion parameter for location *i* across all time points. The priors for the model parameters are defined as follows:
- *β*_*i,t*_ *∼* Gamma(0.001, 0.001),
- 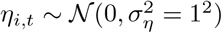,
- *θ*_*i*_ *∼* Gamma(0.001, 0.001),

To ensure consistency across locations and facilitate appropriate prior scaling, each outcome is rescaled to have a maximum of 1000 prior to model fitting. This rescaling is helpful because, across counties, the maximum wastewater values range from 124 to 95,918 and the maximum cases values range from 88 to 279,753. The outcome is then rounded to the nearest integer since the negative binomial distribution can only take integer values. The model is fit using data from all dates on or after the first instance where the outcome exceeds or equals 10 (interpreted as 10 new weekly cases for case data, or 10 RNA copies/mL for wastewater data). This threshold helps the model begin fitting during a more stable period, as very low or zero values tend to increase the risk of divergence in the MCMC sampler.

The model is specified in a Bayesian probabilistic framework and implemented using the `rstan` R package [19], which provides an R interface to Stan [20]. Stan uses the No-U-Turn Sampler (NUTS), an adaptive variant of Hamiltonian Monte Carlo, for efficient Markov Chain Monte Carlo (MCMC) sampling [21]. Each model run uses a burn-in period of 1,000 iterations with 10,000 total samples, which are then thinned by 10 to yield 900 samples after warmup. We use Stan’s default settings of `adapt delta = 0.8` and `max treedepth = 10;` however, if the sampler reports ≥1% divergent transitions, we increase the parameters to `adapt delta = 0.99` and `max treedepth = 12` and rerun the model. Convergence diagnostics for model runs on county wastewater and case data are provided in Section S2.

For each location in the Biobot dataset, we fit the model across all time points [*t*_*i*,0_, *T*_*i*_], where *T*_*i*_ = min (max(*t*_*i*_(cases)), max(*t*_*i*_(wastewater))) is the last date with available cases and wastewater data. The starting point *t*_*i*,0_ is defined as *t*_*i*,first_ + *τ*, where *t*_*i*,first_ is the first time point with wastewater data for location *i* and *τ* = 5 corresponds to a five-week window used to fit the exponential growth model. In other words, *t*_*i*,0_ is the earliest time point at which a full trailing window of length *τ* is available for model fitting. We chose a five-week window to provide six observations for model fitting — enough for stable estimation of exponential growth — while remaining short enough to capture distinct outbreak waves before immunity or behavioral changes alter the trajectory.

Similarly, for each location, we fit the model for reported COVID-19 cases during the same time period [*t*_*i*,0_, *T*_*i*_].

#### 3.1.1 Outbreak Detection Criteria

For each outcome, the model generates posterior estimates for *η*_*i,t*_. From these estimates, we calculate 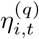, where (q) is the quantile of the posterior, for *q* = 0.05 and *q* = 0.5. We define the start and end of an outbreak as follows:

- An outbreak begins at *t* if 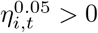 and no outbreak is ongoing.
- An outbreak ends at *t* if 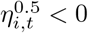 while an outbreak is active.

At any given time, only one outbreak can be active for a location *i*. An outbreak start is defined only if there is no ongoing outbreak, and an outbreak end is defined only if there is an active outbreak. This ensures that outbreaks are non-overlapping and sequential within each location.

We chose the threshold 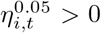 to mark the start of an outbreak to ensure high posterior certainty (at least 95%) that there is exponential growth at time *t*. To define the end of an outbreak, we apply the more lenient condition 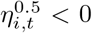, which reflects the model is uncertain (posterior probability *<* 50%) about the continuation of exponential growth. We chose this relaxed criterion because we do not require certainty about exponential decay - only that the model is no longer certain that exponential growth is ongoing.

### 3.2 Peak Detection Approach

While the onset detection approach captures outbreak onsets as periods of sustained positive growth, a complementary objective is identifying epidemic peaks—the time points at which wastewater or case signals reach local maxima. Peak detection provides a distinct and interpretable signal of epidemic dynamics, enabling evaluation of whether wastewater surveillance can track the trajectory of an outbreak through its apex. Because peak timing is often operationally relevant and because a simple signal-based detector may be sufficient for this task, we develop a rule-based peak detection approach that complements the onset detection framework while remaining easy to interpret and implement.

A peak is detected at time *t*_*p*_ with the following rules for *y* ∈ {wastewater, cases, hospitalizations, deaths}:

- *y*(*t*_*p*_) *> y*(*t*) for *t* ∈ [*t*_*p*_ *− w*_*L*_, *t*_*p*_)
- *y*(*t*_*p*_) *≥ y*(*t*) for *t* ∈ (*t*_*p*_, *t*_*p*_ + *w*_*R*_]
- *y*(*t*_*p*_) *>* 10
- *y*(*t*) ≠ 0 for all *t* ∈ [*t*_*p*_ *− w*_*L*_, *t*_*p*_ + *w*_*R*_] that *y*(*t*) is observed

where *w*_*L*_ and *w*_*R*_ are the left and right window lengths. The strict inequality for the values on the left ensures that for repeating values (e.g. two weeks in a row with 100 cases), the first value is detected as the peak while the second is not. Note that the *y*(*t*_*p*_) *>* 10 threshold was used for each data source. This was appropriate for filtering out only the smallest points within the range of each data source in this study; however, for other data sources this might need to be adjusted to properly reflect the magnitude.

For the main results in this study, we set *w*_*L*_ = *w*_*R*_ = 3. Other rules for peak detection—such as different window lengths and signal-to-noise thresholds—are tested in Appendix Section S3. None of these rules consistently improved peak detection results by fitting on wastewater and cases, so we used the simple rule based approach described here.

### 3.3 Curated Outcomes

#### 3.3.1 Curated Case Outbreaks

While using *exp growth* to define case-based outbreaks enables internal validation, the resulting reference events reflect the model’s particular assumptions and parameterization rather than an independent epidemiological judgment of outbreak timing. Curated case outbreaks address this by substituting expert review for model-defined labels, providing a reference outcome that is not beholden to the same algorithmic choices.

This curated outcome begins with the set of outbreaks detected by the onset detection approach applied to reported case data. Each outbreak was then visually inspected using plots of the case time series. Outbreaks were either removed (e.g., if they appeared implausible or reflected short-term noise), merged (e.g., if two nearby outbreaks were judged to represent a single sustained event), date-adjusted (i.e. shifting starting or ending time), or added. Very few new outbreaks were added: visual review did not identify many missed outbreaks that the model failed to detect—in fact, none of the county-aggregated or state-level results yielded added outbreaks—only the county results did. At the county-level, out of 421 algorithmic outbreaks, 140 were removed, 43 had their end date changed, and 26 were added.

The curated outbreaks reflect both the structure of the exponential growth model and the expert judgment of a human reviewer. This outcome is used alongside purely model-defined outbreaks to evaluate wastewater-based detection performance.

The curated outbreaks were applied to all county-aggregated and state-level data; however, for the county data the curation was restricted to counties with population of at least 100,000 on the 2020 census, leaving 184 out of 281 counties in the dataset. Counties with smaller populations have noisier case time series and it is more ambiguous for a human to define outbreaks from them.

#### 3.3.2 Curated Peaks

An analogous curation process was applied to peaks detected by the rule-based peak detection approach. Beginning with the algorithmically detected peaks in the case time series, each peak was visually inspected and assessed for plausibility. Unlike the outbreak curation, peaks were only removed during this process—no peaks were added and no peak times were shifted. A substantial proportion of algorithmically detected peaks were removed upon review, as many reflected noise, reporting artifacts, or minor fluctuations that did not correspond to meaningful epidemic peaks. Out of 2133 algorithmically detected peaks at the county-level, 804 were removed. The curated peaks serve as the reference outcome for evaluating peak detection performance in wastewater data.

Supplementary Section S1 reports inter-rater agreement between two independent reviewers on curated outbreak and peak definitions.

### 3.4 Outbreak and Peak Classification

The classification framework described here applies to both the onset detection approach (Section 3.1) and the peak detection approach (Section 3.2), with minor differences noted where applicable. This framework is a central methodological contribution of the paper: it translates wastewater surveillance into an event-detection evaluation problem and makes explicit the assumptions required to compare wastewater signals to imperfect clinical reference outcomes.

#### 3.4.1 Classification Definitions

To evaluate the ability of wastewater signals to identify events also found in clinical outcome data, we define a **detection interval** around each outcome event:

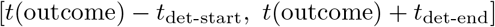

where *t*(outcome) is the onset of an outcome outbreak or the time of an outcome peak. A wastewater event is counted as a match if it occurs within this interval. Thus, the detection interval defines how early or late a wastewater signal may occur and still be considered a valid detection of the corresponding clinical event.

For comparisons with cases, we use a symmetric detection interval of [−3, 3] weeks for both outbreaks and peaks, allowing wastewater to lead or lag the case-defined event by up to three weeks. For comparisons with hospitalizations and deaths in aggregated county-level and state-level analyses, we use detection intervals of [−5, 1] and [−6, 0], respectively, reflecting the longer delays from infection to those outcomes. Although these centered lags are only rough approximations and do not account for delays between infection and wastewater signal onset or case reporting, the six-week intervals offer sufficient flexibility to capture timing variability across indicators and outcomes.

We define the following classification types:

##### Definitions

- **True Positive (TP)**: A wastewater event occurs at time *t*(wastewater) and an outcome event occurs at *t*(outcome), such that *t*(wastewater) falls within the detection interval for *t*(outcome):

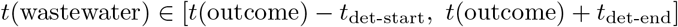
- **False Positive (FP)**: A wastewater event occurs at time *t*(wastewater) without any *t*(outcome) such that *t*(wastewater) falls within the corresponding detection interval. For outbreaks, no active outcome outbreak may be ongoing.
- **False Negative (FN)**: An outcome event occurs at *t*(outcome) without any *t*(wastewater) falling within the corresponding detection interval. For outbreaks, no active wastewater outbreak may be ongoing at *t*(outcome).

We only classify outbreak TPs, FPs and FNs when no outbreak is already ongoing in the corresponding data stream, to avoid penalizing the model for continued detection of a persistent outbreak. This restriction does not apply to peak detection, as peaks are instantaneous events with no defined duration. We do not define true negatives due to the difficulty of unambiguously characterizing non-event periods given overlapping detection windows.

##### Data Completeness

To avoid scoring events when wastewater coverage is too sparse to allow reliable detection, we exclude classifications where less than two-thirds of the weeks in the relevant wastewater window have observed measurements.

For **outbreak** TP and FP classifications, this window corresponds to the exponential growth model fitting window ending at the wastewater outbreak start:

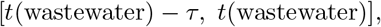

with *τ* = 5 weeks. For outbreak FN classifications, the window spans the entire period in which a wastewater outbreak could have been detected, plus the fitting window:

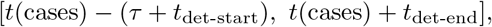

where *t*_det-start_ = *t*_det-end_ = 3, yielding a window of [−8, +3] weeks relative to the case outbreak start.

For **peak** classifications, data completeness is assessed over the detection window centered on the peak. For peak TP and FP classifications, this window is:

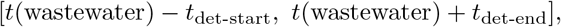

where *t*(wastewater) is the detected wastewater peak time. For peak FN classifications, coverage is assessed over the broader window that spans both the peak detection window and the detection interval:

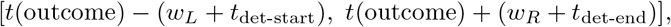

where *w*_*L*_ and *w*_*R*_ are the left and right window lengths used in the peak detection approach. In all cases, classifications are excluded when observed wastewater completeness falls below two-thirds.

##### Censoring

Classifying events near the start or end of the time series can introduce bias due to incomplete data availability. For example, an outcome outbreak occurring at the beginning of the time series may be misclassified as a false negative if a corresponding wastewater signal would have been detected earlier, had data collection started sooner.

To address this, we restrict classification to events where the full detection and fitting windows are contained within the observed time series. For **outbreak** classifications:

- TPs and FNs are restricted to *t*(cases) *∈* [*t*_*i*,0_ + *τ* + *t*_det-start_, *T*_*i*_ *− t*_det-end_]
- FPs are restricted to *t*(wastewater) *∈* [*t*_*i*,0_ + *τ* + *t*_det-end_, *T*_*i*_ *− t*_det-start_]

For **peak** classifications:

- TPs and FNs are restricted to *t*(cases) *∈* [*t*_*i*,0_ + *w*_*L*_ + *t*_det-start_, *T*_*i*_ *− w*_*R*_ *− t*_det-end_].
- TPs (as wastewater peak date) and FPs are restricted to *t*(wastewater) *∈* [*t*_*i*,0_+*w*_*L*_+*t*_det-end_, *T*_*i*_*−w*_*R*_*−t*_det-start_].

These restrictions ensure that each event has a valid opportunity for detection. While they exclude a subset of events at the boundaries of the time series and reduce sample size, they improve the validity of our classification metrics.

Because the censoring boundaries depend on the detection interval parameters *t*_det-start_ and *t*_det-end_, the evaluable time windows differ across outcomes. Specifically, comparisons against deaths censor more events at the end of the time series, while comparisons against cases apply a symmetric window. As a result, performance metrics for different outcomes are evaluated over slightly different calendar periods, which should be kept in mind when making direct cross-outcome comparisons.

##### Metrics

We calculate the following metrics across all locations and within each location:

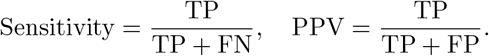

Sensitivity signifies the proportion of clinical events that are detected by wastewater events and positive predictive value (PPV) signifies the proportion of wastewater events that are true alarms. These metrics are computed separately for outbreak detection and peak detection.

### 3.5 County Aggregated and State-Level Comparison with Hospitalizations and Deaths

COVID-19 hospitalizations and deaths have been widely used to monitor epidemic trends. They are considered relatively stable indicators of disease trends, particularly in the absence of case reporting [22]. Therefore, we assess the performance of wastewater outbreak detection against these outcomes in addition to cases. However, both data availability and signal stability present limitations for using hospitalizations and deaths at the county-level.

The hospitalization data available uniformly across the US are only available at the state-level. While death data are available at the county-level, the counts are generally too low to yield stable estimates of exponential growth. Consequently, we restrict analyses involving hospitalization to the state-level and deaths to the county-aggregated and state-level.

Beyond data availability, comparing outbreak detection performance between the county- and state-level aggregation is informative. Aggregating data across larger geographic areas can improve the signal-to-noise ratio of both wastewater and cases and enable more accurate detection due to smoother time series. However, this may also mask important heterogeneity and limit the ability to detect localized outbreaks.

We fit the exponential growth model on all data sources at multiple spatial resolutions. All data sources are restricted to the period from January 6, 2021 (the earliest date with wastewater data) to March 8, 2023 (the last date with case data) to enable valid cross-series comparisons. While cases, hospitalization, and death data are complete over this interval, wastewater data are sparse and inconsistent across states due to logistical constraints in sampling. We assume that the incomplete data are missing completely at random with respect to outbreak dynamics as these are the result of administrative decisions, and therefore do not bias the results systematically.

Different indicator-outcome pairs are expected to exhibit characteristic time lags. The expected delay from symptom onset to first clinical visit is 5 days, to ICU admission is 10 days, and to death is 16 days [23]. These expected delays motivate the detection intervals described in Section 3.4, with wider pre-outcome windows used for hospitalizations and deaths than for cases.

## 4 Results

Using the classification framework introduced above, we evaluate wastewater as an event-detection system for outbreak onset and epidemic peak timing. We find that the onset detection approach provides strong outbreak detection performance, outperforming a benchmark *R*_*t*_-based method (Appendix Section S2), while the rule-based peak detection approach also performs well across settings. The results suggest that wastewater can often identify epidemiologically meaningful events in a timely way, but that measured performance depends on the event definition, the reference clinical outcome, and the spatial scale of analysis. Overall, the empirical findings support the broader framing of wastewater surveillance as a practical event-detection tool rather than only as a source for correlation analysis or forecasting.

### 4.1 Outbreak Detection Performance

Figure 1 shows exponential model fits across multiple time windows for Los Angeles (LA) County. The fitted curves closely follow the observed data, and the posterior predictive intervals appropriately capture uncertainty. Notably, the model shows high confidence in exponential growth during the third time window (ending 11/30/2022).

**Figure 1:**
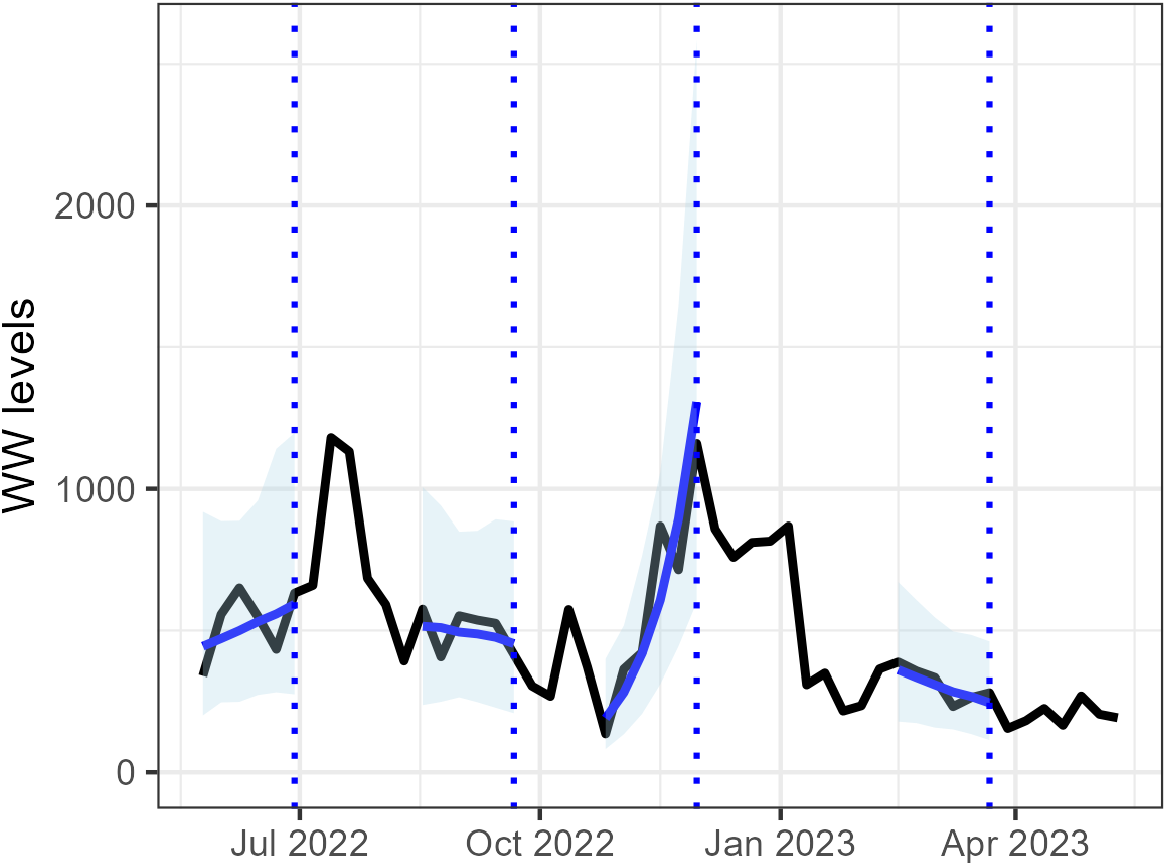
Exponential model fit on Los Angeles county showing prediction windows where the model is fit on 6/29/2022, 9/21/2022, 11/30/2022, and 3/22/2023 (shown as vertical dashed lines). The observed data is in black. The solid blue line shows the median posterior prediction and the shaded blue region shows the 95% posterior predictive interval.

Figure 2 displays classified outbreak detections in LA using the onset detection approach on both wastewater and cases. The wastewater onset detection approach correctly identified two case-based outbreaks on 7/20/2022 and 11/16/2022 (orange triangles).

**Figure 2:**
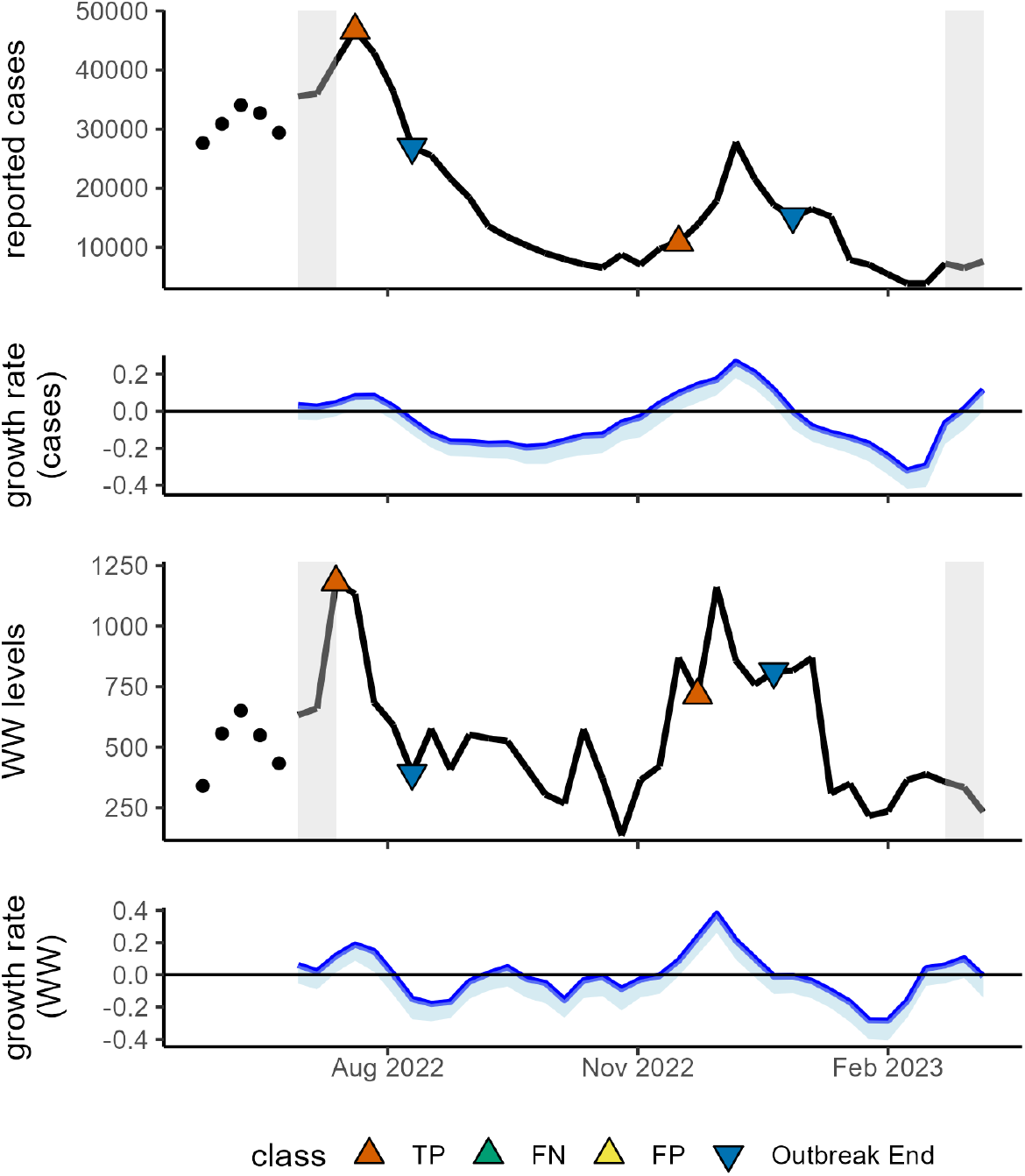
Wastewater and cases outbreak detections and classifications for Los Angeles County. The top panel shows reported cases, with outbreak onsets marked by upwards triangles and endings by downwards triangles. The second panel displays estimates of eta (the growth parameter) from the cases exponential growth model, with the solid blue line representing the median posterior estimate and the shaded blue region representing the (5%, 50%) posterior credible interval. The third and fourth panels show the reported levels and growth rates for the wastewater data, respectively. Orange, green, and yellow colors indicate true positive, false negative, and false positive outbreak classifications. The shaded gray bars represent censoring regions where classifications are excluded.

**Figure 3:**
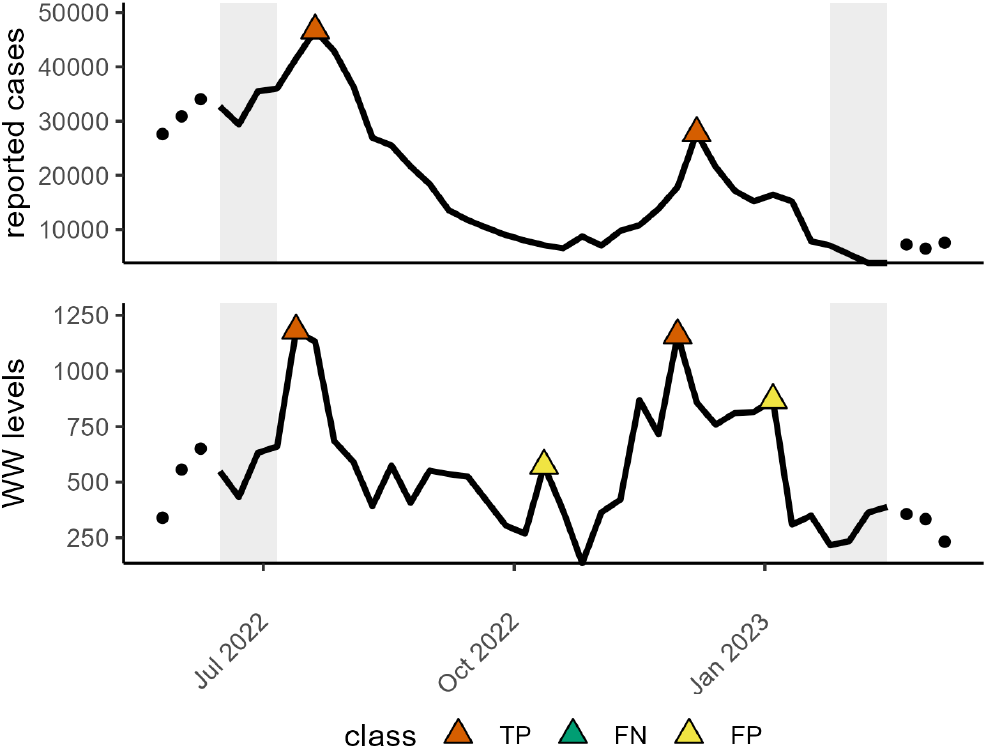
Peak detection results for Los Angeles County. The top panel shows the wastewater time series with detected wastewater peaks marked by filled triangles. The bottom panel shows the reported case time series with detected case peaks marked by filled triangles. Orange triangles indicate true positive detections—wastewater peaks that fall within the detection interval of a corresponding case peak—while green triangles indicate false negative case peaks not detected by wastewater and yellow triangles indicate false positive wastewater peaks with no matching case peak. The shaded gray bars represent censoring regions where classifications are excluded.

Table 1 summarizes outbreak detection performance across all spatial scales and clinical outcomes using the onset detection approach. At the county level, wastewater identifies about four out of five case-defined outbreaks, with about two out of three wastewater detections being correct (sensitivity 0.82, PPV 0.64). Against curated case outbreaks, sensitivity rises to 0.92, suggesting that wastewater detections often align with outbreaks deemed visually plausible by expert curation; the lower PPV (0.57) reflects that some events flagged by wastewater but excluded during curation may still reflect real outbreak signals, particularly where case data are sparse or ambiguous. Mean wastewater outbreak lead times of 0.62–0.83 weeks (4.3 - 5.8 days) across spatial scales suggest a modest but potentially useful lead time for operational event detection, in line with previous research. Detailed lead time distributions are provided in Appendix Section S4, which show that most wastewater detections precede or coincide with case outbreaks within a 0–2 week lead window.

**Table 1:** Outbreak detection results across different dataset combinations using the onset detection approach. Detection interval is the allotted window in weeks where a wastewater outbreak is treated as a true positive of the outcome: [−3, 3] for cases, [−5, 1] for hospitalizations, and [−6, 0] for deaths. Curated cases are only available for counties with population of at least 100,000. Mean Lead is the average number of weeks by which the wastewater outbreak precedes the outcome outbreak across all true positives. Positive values indicate the wastewater outbreak occurs first. Full method comparisons including the *deconv-Rt* baseline are provided in Appendix Section S2.

| Aggregation | Outcome | Detection Interval | Sensitivity | PPV | TPs | FPs | FNs | Mean Lead |
| --- | --- | --- | --- | --- | --- | --- | --- | --- |
| county | cases | $[-3, 3]$ | 0.82 | 0.64 | 306 | 175 | 67 | 0.62 |
| county ($\geq 100,000$ population) | cases | $[-3, 3]$ | 0.88 | 0.69 | 240 | 109 | 33 | 0.68 |
| county ($\geq 100,000$ population) | curated cases | $[-3, 3]$ | 0.92 | 0.57 | 196 | 147 | 17 | 0.72 |
| county-aggregated | cases | $[-3, 3]$ | 0.88 | 0.71 | 99 | 41 | 14 | 0.64 |
| county-aggregated | curated cases | $[-3, 3]$ | 0.92 | 0.61 | 85 | 55 | 7 | 0.69 |
| county-aggregated | deaths | $[-6, 0]$ | 0.60 | 0.41 | 52 | 75 | 34 | 3.52 |
| state | cases | $[-3, 3]$ | 0.87 | 0.69 | 94 | 43 | 14 | 0.81 |
| state | curated cases | $[-3, 3]$ | 0.94 | 0.62 | 83 | 51 | 5 | 0.83 |
| state | hospitalizations | $[-5, 1]$ | 0.91 | 0.75 | 94 | 31 | 9 | 2.20 |
| state | deaths | $[-6, 0]$ | 0.73 | 0.79 | 91 | 24 | 34 | 2.08 |

There is limited evidence that wastewater-based outbreak detection improves at higher spatial aggregations, but none of the comparisons are statistically significant at the *α* = 0.05 level. Sensitivity for the cases outcome increases from 0.82 at the county-level to 0.88 for county-aggregated data (p = 0.16 from a two-proportion z-test) and 0.87 for state-level data (p = 0.22). PPV also changes from 0.69 at the county-level to 0.71 (p = 0.12) and 0.69 (p = 0.28), respectively. Using curated cases, sensitivity remains relatively constant across spatial aggregations while PPV increases: sensitivity changes from 0.92 for counties with at least 100,000 population to 0.92 (p = 0.91) and 0.94 (p = 0.49), while PPV increases from 0.57 to 0.61 (p = 0.47) and 0.62 (p = 0.34). These patterns are consistent with modest benefits of spatial aggregation for outbreak detection stability, but the evidence is not strong enough here to conclude a clear aggregation effect. Because county-aggregated and state-level case data have similar results, any benefit of additional aggregation appears limited in these data.

Wastewater performs better when detecting deaths at the state-level (sensitivity 0.73, PPV 0.79) than at the county-aggregated level (sensitivity 0.6, PPV 0.41). This aligns with prior work showing that sparse death counts at finer geographic scales limit information for epidemiological analysis [24]. Aggregation likely improves signal quality by smoothing random variation in sparse data.

Among the state-level clinical outcomes, wastewater achieves the highest sensitivity when detecting curated case outbreaks and the highest PPV when detecting deaths. However, its overall performance is relatively strong across all three outcomes: cases, hospitalizations, and deaths. Note that because censoring boundaries depend on the outcome-specific detection intervals, the evaluable time windows differ slightly across outcomes. Because wastewater is subject to different sources of bias and confounding than clinical outcomes, these alignments likely reflect a genuine signal of underlying transmission dynamics. Appendix Section S5 further examines false positives and false negatives, showing that many misclassified events exhibit partial signal in the opposing data stream, suggesting they may reflect near-threshold detections rather than true errors. The gap between algorithmic and curated PPV — particularly for peak detection, where county-level PPV drops from 0.68 to 0.40 — suggests that a substantial fraction of apparent false positives reflect the noisiness of the case reference standard rather than wastewater detection errors, and that PPV estimates should be interpreted accordingly.

Taken together, these findings suggest that outbreak detection using wastewater is effective at multiple spatial scales. The results are compatible with modest benefits of aggregation in some settings, particularly for noisier outcomes, but they do not show a clear or statistically significant aggregation effect across all comparisons. These findings reinforce the utility of WBS for outbreak detection at multiple levels of public health infrastructure, from local to state-wide systems.

From an epidemiological perspective, the observed lead times for wastewater-based outbreak detection are consistent with infectious disease transmission dynamics. Because viral shedding into wastewater occurs soon after infection and can precede symptom onset, testing, and case reporting, wastewater captures this growth phase closer to the time of infection than clinical surveillance does. The modest lead times observed here (approximately 0.6–0.8 weeks for cases) therefore suggest that wastewater functions less as a long-range forecasting tool than as a near-real-time indicator of underlying transmission. The longer lead times relative to hospitalizations and deaths are also epidemiologically consistent, since those outcomes occur farther downstream along the infection-to-outcome pathway.

Finally, the onset detection approach substantially outperformed the Rt-based baseline. When applied to both wastewater and cases, the onset detection approach achieves sensitivity 0.82 and PPV 0.64, compared with sensitivity 0.58 and PPV 0.19 for the best-performing Rt variant (deconv-Rt(0.25)); full method comparisons are provided in Appendix Section S2. This performance gap likely reflects the simpler, single-stage structure of the exponential growth model, which avoids the additional uncertainty introduced by deconvolution and Rt estimation steps and better accommodates overdispersed data through a negative binomial likelihood.

### 4.2 Peak Detection Performance

Table 2 presents peak detection performance across spatial aggregations and clinical outcomes. Overall, the peak detection approach demonstrates strong and consistent performance. When using algorithmic cases as the reference outcome, sensitivity and PPV are nearly identical at the county (0.84, 0.70), county-aggregated (0.81, 0.68), and state levels (0.85, 0.70), indicating that spatial aggregation has little effect on peak detection performance against this reference standard. However, results using curated cases tell a different story: sensitivity is stable across spatial scales (0.88, 0.90, and 0.92 at the county, county-aggregated, and state levels; p = 0.63 and p = 0.27 comparing county-aggregated and state to county, respectively), while PPV increases significantly with aggregation (0.40, 0.48, and 0.52; p = 0.014 and p = 0.0005).

**Table 2:**
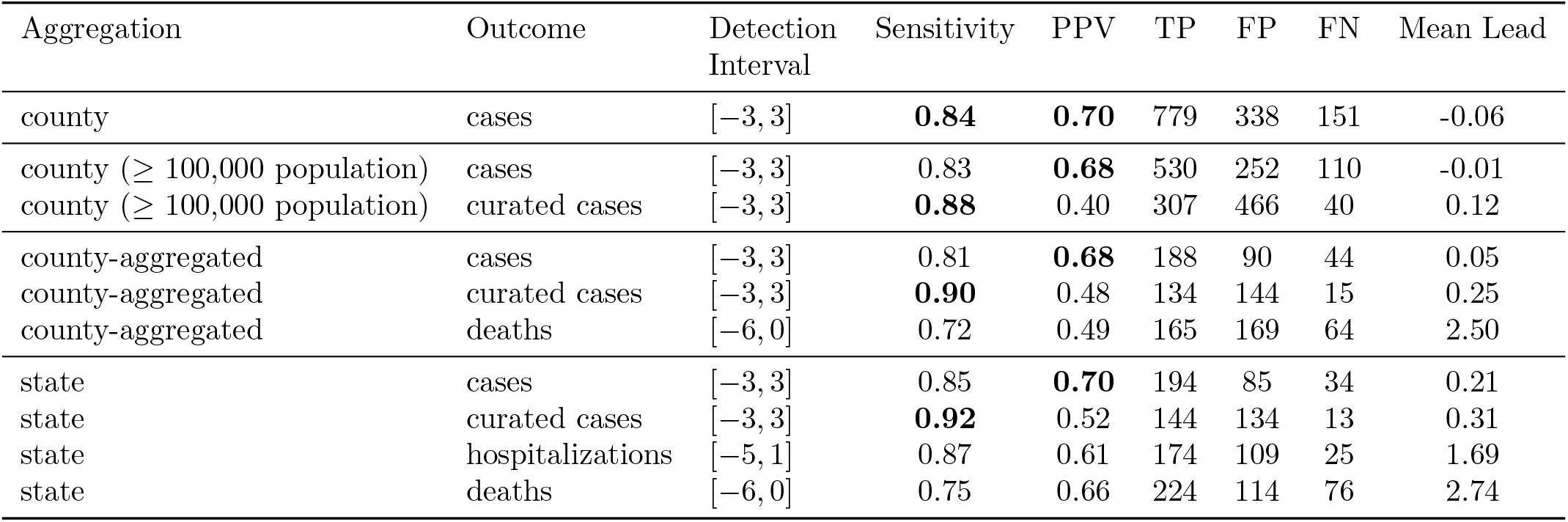
Peak detection results across different dataset combinations. The peak detection approach is applied to both wastewater and outcome data (cases, curated cases, hospitalizations, or deaths). Detection interval is the allotted window in weeks where a wastewater outbreak is treated as a true positive of the outcome: [− 3, 3] for cases, [−5, 1] for hospitalizations, and [− 6, 0] for deaths. Mean Lead is the average number of weeks by which the wastewater peak precedes the outcome peak across all true positives (negative values indicate the wastewater peak followed the outcome peak on average). All methods use a peak detection window of *w* = 3.

| Aggregation | Outcome | Detection Interval | Sensitivity | PPV | TP | FP | FN | Mean Lead |
| --- | --- | --- | --- | --- | --- | --- | --- | --- |
| county | cases | $[-3, 3]$ | **0.84** | **0.70** | 779 | 338 | 151 | -0.06 |
| county ($\geq 100,000$ population) | cases | $[-3, 3]$ | 0.83 | **0.68** | 530 | 252 | 110 | -0.01 |
| county ($\geq 100,000$ population) | curated cases | $[-3, 3]$ | **0.88** | 0.40 | 307 | 466 | 40 | 0.12 |
| county-aggregated | cases | $[-3, 3]$ | 0.81 | **0.68** | 188 | 90 | 44 | 0.05 |
| county-aggregated | curated cases | $[-3, 3]$ | **0.90** | 0.48 | 134 | 144 | 15 | 0.25 |
| county-aggregated | deaths | $[-6, 0]$ | 0.72 | 0.49 | 165 | 169 | 64 | 2.50 |
| state | cases | $[-3, 3]$ | 0.85 | **0.70** | 194 | 85 | 34 | 0.21 |
| state | curated cases | $[-3, 3]$ | **0.92** | 0.52 | 144 | 134 | 13 | 0.31 |
| state | hospitalizations | $[-5, 1]$ | 0.87 | 0.61 | 174 | 109 | 25 | 1.69 |
| state | deaths | $[-6, 0]$ | 0.75 | 0.66 | 224 | 114 | 76 | 2.74 |

Peak detection performance varies more by outcome type than by spatial scale for algorithmic outcomes. At the state level, sensitivity against hospitalizations is the highest among all algorithmic outcomes (0.87), but PPV is lower (0.61) compared to cases (0.70) and deaths (0.66).

Deaths show lower sensitivity and PPV than cases at both the county-aggregated and state levels, but exhibit a notable improvement in PPV from the county-aggregated level (0.49) to the state level (0.66), mirroring the pattern seen in outbreak detection. This improvement again points to the benefit of aggregation for the noisiest and most delayed outcomes.

Using curated cases as the reference outcome reveals a notable divergence from the algorithmic cases results: sensitivity is largely stable across scales while PPV is substantially lower at all aggregation levels (0.40, 0.48, and 0.52 compared to 0.68–0.70 for algorithmic cases). This pattern reflects the fact that curated cases apply a stricter standard for what constitutes a meaningful epidemic peak than the algorithmic peak detection approach. As a result, many algorithmic case peaks that were aligned with wastewater peaks—and therefore counted as TPs against algorithmic cases—are excluded from the curated reference, reclassifying those wastewater detections as FPs. The significant improvement in curated PPV with aggregation (but not in algorithmic PPV) further suggests that many of these reclassified FPs reflect genuine transmission signals that become visually recognizable in smoother, aggregated case series.

Notably, the mean lead time between wastewater and case peaks is near zero across all spatial scales (ranging from -0.06 to 0.31 weeks across algorithmic and curated case outcomes), in contrast to outbreak detection where wastewater consistently leads cases by 0.6–0.8 weeks. This could reflect that viral shedding often begins before cases are recorded, while shedding may continue after case identification. Lead times are substantially larger for hospitalizations (1.69 weeks) and deaths (2.50–2.74 weeks), consistent with the expected biological delays between infection, clinical presentation, and death. Detailed county-level lead time distributions are provided in Appendix Section S4, showing a wide spread of peak lead times between -3 and 3.

Together, these findings suggest that wastewater-based peak detection is robust across a range of outcomes and spatial scales. Performance improves with spatial aggregation when a stricter curated reference standard is applied, suggesting a clearer visual signal for expert reviewers rather than improved wastewater detection at higher spatial scales. In the broader framing of the paper, this is important because peak timing is often a simpler and more attainable event-detection target than forecasting full epidemic trajectories, while still being operationally meaningful. The rule-based peak detection approach performs comparably to the more complex onset detection approach, and its performance against algorithmic outcomes is not strongly dependent on spatial aggregation. This makes peak detection a practical complement to outbreak detection for monitoring epidemic trajectories in near real-time.

The near-zero lead time between wastewater and case peaks can be understood through the distinction between incidence and prevalence. Reported cases approximate incidence, capturing new infections at a single point in time, whereas wastewater reflects a form of prevalence, aggregating viral shedding over the course of an individual’s infectious period. Because infected individuals can shed virus for many days after infection—often more than a week—wastewater signals include contributions from both recent and earlier infections. During the early phase of an outbreak, when most infections are new and exponential growth dominates, wastewater can lead case data by capturing infections closer to their time of occurrence. In contrast, near the epidemic peak, wastewater reflects a mixture of newly infected individuals and those infected during the growth phase who continue shedding. This accumulation of “trailing” viral shedding dampens temporal differences between wastewater and incidence signals, helping explain the observed synchronization of peak timing.

## 5 Discussion

This study contributes to the wastewater surveillance literature in three ways. First, it frames wastewater surveillance as an event-detection problem focused on outbreak onset and epidemic peak timing rather than on correlation or prediction alone. Second, it develops a classification-based framework for evaluating how well wastewater signals detect epidemiologically meaningful events under explicit assumptions about timing, completeness, censoring, and imperfect reference outcomes. Third, it provides an empirical assessment of outbreak and peak detection across approaches, spatial scales, and clinical outcomes. Within this framework, both the onset detection and peak detection approaches show that wastewater can provide practically useful signals of epidemic dynamics, although measured performance depends on the event definition, the reference outcome, and the spatial scale of analysis.

Unlike many prior studies that focus on correlation or forecasting, our work is motivated by how surveillance systems are often used in practice: to determine whether an outbreak is beginning, whether transmission is accelerating, or whether an epidemic has likely peaked. Forecasting continuous incidence trajectories is a distinct and more demanding task; state-of-the-art forecasting models have repeatedly struggled to anticipate sharp increases in disease activity in real-time operational settings, even when performing well retrospectively [11]. This event-based perspective does not replace forecasting, which remains valuable for planning and scenario analysis, but it emphasizes a complementary and often more attainable target for surveillance. In noisy real-world settings, accurate forecasting can be difficult, whereas reliable identification of outbreak onset or peak timing may be both more feasible and more directly actionable. The classification framework developed here is intended to support that practical use case by making explicit what counts as successful detection, what timing mismatches are allowed, and how false alarms and missed detections should be interpreted.

A key contribution of the study is therefore not only the use of the onset detection and peak detection approaches, but the explicit evaluation framework used to compare wastewater against clinical outcomes. By defining detection intervals, classification rules, censoring restrictions, and completeness criteria, we translate wastewater surveillance into an event-detection evaluation problem that can be compared across settings and outcomes. This is analogous in spirit to forecast evaluation frameworks that rely on standardized outcomes and metrics, but it is adapted to the more operational question of whether a surveillance signal successfully detects meaningful epidemiological events. Because outbreak and peak detection are often how surveillance signals are used by public health authorities, we view this framework as a practically grounded contribution that may generalize beyond COVID-19 and beyond wastewater.

Peak detection, which requires no model fitting, performed comparably to the onset detection approach overall, with county-level sensitivity 0.84 and PPV 0.70 against algorithmic case peaks. The onset detection performed well at the county level, substantially outperforming the Rt-based baseline (sensitivity 0.82 and PPV 0.64 versus 0.58 and 0.19), consistent with the simpler, single-stage structure of the exponential growth model avoiding the compounded uncertainty of deconvolution and Rt estimation. Both methods demonstrated strong performance against hospitalizations and deaths at the state level. Performance against deaths was weaker at finer spatial scales, reflecting low counts and high variability in county-level death data. Spatial aggregation showed modest, statistically non-significant improvements for outbreak detection, and stable performance for peak detection against algorithmic case outcomes; PPV for peak detection improved significantly with aggregation only against the curated reference standard. Appendix Section S6 further explores how detection performance varies with population size and sampling intensity, showing that outbreak detection is generally stronger in larger counties while peak detection remains stable across settings. The near-zero lead time between wastewater and case peaks contrasts with outbreak detection, where wastewater consistently precedes cases by 0.6–0.8 weeks. Lead times for hospitalizations and deaths are consistent with expected biological delays: for outbreak detection, wastewater leads hospitalizations by 2.2 weeks and deaths by 2.1–3.5 weeks; for peak detection, the corresponding leads are 1.7 and 2.5–2.7 weeks. These patterns further support the interpretation that wastewater captures upstream transmission dynamics regardless of detection approach.

The results can also be interpreted through the lens of epidemic theory and the distinction between incidence and prevalence. In compartmental models such as SIR, the early phase of an epidemic is characterized by approximately exponential growth when *R*_*t*_ > 1, making exponential growth a mechanistically grounded approximation of early transmission dynamics. Reported cases approximate incidence, capturing new infections at a single point in time, whereas wastewater reflects a form of prevalence by aggregating viral shedding over the infectious period, which extends beyond the point of infection captured by incidence-based measures. This makes wastewater particularly sensitive to the accumulation of recent infections during the growth phase, while case data are subject to delays from testing and reporting. The strong performance of the onset detection approach for outbreak detection is therefore consistent with the expectation that early exponential transmission is more directly observable in wastewater signals than in case data. However, because wastewater reflects accumulated prevalence rather than point incidence, it does not consistently lead case data in peak detection when both recent and past infections contribute to the signal. The classification framework in this study enables these distinctions to be evaluated explicitly, separating conditions under which wastewater acts as a leading indicator from those in which it does not.

There are several limitations to our study. First, we treat case-based events as the ground truth when evaluating wastewater-based detection, even though case data are a biased and noisy proxy for true infections. As a result, our classification labels (true positives, false positives, and false negatives) are subject to label noise: some events identified as false positives in wastewater may reflect true transmission signals that are not captured in case data, while some missed detections may arise from errors or delays in case reporting (such as backlogs of confirmed cases being reported all in one week [25, 26]). If errors in wastewater and case data are independent, our estimates likely underestimate the true performance of wastewater-based detection, as the reference outcome is itself imprecise. If the errors are positively correlated, however, the opposite may be true. There is no clear mechanism by which these errors would be systematically correlated; one strength of wastewater-based surveillance is that it operates independently of clinical testing and reporting, suggesting that our performance estimates are likely conservative.

The manually curated case events were introduced precisely to address some of these reference standard limitations, but curation introduces its own constraints. While curated outcomes helped correct implausible algorithmic detections, they still rely on observed case data and introduce an additional layer of subjectivity. For outbreaks, the divergence between model-defined and expert-curated results underscores a broader challenge: statistically detected periods of exponential growth do not always align with visually or operationally defined outbreaks. For peak detection, this discrepancy was even more pronounced—the sharp drop in PPV when using curated rather than algorithmically defined case peaks reflects the stricter standard applied during curation, which excludes many minor fluctuations that the rule-based peak detection approach treats as peaks. Inter-rater agreement between two independent reviewers was high for both outbreaks and peaks (Supplementary Section S1), though residual disagreement confirms that expert curation of these events is itself subject to some subjectivity.

More broadly, results depend on the specific definitions used to identify outbreaks and peaks and to match events across data streams. These include the choice of detection interval (e.g., ±3 weeks for cases), the thresholds used to define outbreak start and end in the onset detection approach, and the rules used for peak detection. Different choices—such as narrower or wider detection intervals, alternative growth thresholds, or different peak definitions—could lead to different classifications and therefore different estimates of sensitivity and PPV. While we selected these definitions to reflect plausible epidemiological timing and to maintain consistency across analyses, there is no universally accepted standard for defining outbreak onset, peak timing, or event alignment across data sources. Together, these considerations highlight a fundamental limitation of any event-based evaluation: both the reference labels and the event definitions influence measured performance, and results should be interpreted in that context.

Beyond the evaluation framework, the modeling approach itself has limitations. Even simpler models than the exponential growth model—such as a frequentist regression of log-transformed values—could yield comparable or better performance for exponential growth detection. The outcome distribution for the exponential growth model (negative binomial) required rounding wastewater data, which could bias results in low incidence periods; however, wastewater was scaled to be in a large range (1 to 1000) before rounding, so this likely has little impact. Different outcome distributions could be used. Additionally, a hierarchical Bayesian model that borrows information across space and time could improve the stability of the onset detection approach. The five-week model fitting window may delay outbreak detection, and this delay would not be identified in this study since wastewater and clinical detections are based on the same approach. Future work should assess detection timing more directly, particularly in contexts with more frequent sampling that could allow for fitting the model on shorter time scales.

Additional limitations include our spatial aggregation strategy, which treated all states equally despite large differences in state size, geographic spread of wastewater sampling sites, and number of sampling sites—differences that may influence the benefits of aggregation. The z-tests comparing outbreak and peak detection sensitivity and PPV across spatial scales were not corrected for multiple comparisons and should be interpreted as exploratory rather than confirmatory. More broadly, the classification framework proposed here can be extended and refined in future work, for example by examining alternative event definitions, alternative reference outcomes, or probabilistic rather than binary evaluation rules. We view those extensions as a natural next step in developing a more general science of wastewater-based event detection.

Wastewater-based surveillance is being implemented worldwide to track a wide range of infectious diseases, including cholera, mpox, influenza, typhoid, and antimicrobial resistance [2]. Our findings demonstrate that wastewater data can reliably detect COVID-19 outbreaks and track epidemic peaks, suggesting its potential for broader pathogen surveillance. More generally, the study supports a shift in emphasis from asking only whether wastewater correlates with clinical outcomes or improves forecasts to asking whether it can reliably detect the epidemiological events that matter for decision-making. Performance is not uniform across locations or spatial scales, but these results support wastewater surveillance as a useful tool for detecting outbreaks and peaks in conjunction with other public health data streams.

## Data Availability

The code used to process the data, fit the models, and reproduce the analyses in this study is available from the authors upon reasonable request. The wastewater data used in this study were provided by a commercial data provider and are not publicly available. Restrictions on redistribution prevent the public release of these data.

## Acknowledgements

We thank Biobot Analytics for providing the wastewater data used in this analysis. We thank Mariana Matus and Joshua Harrison for meaningful scientific discussions that helped us understand the data and that informed the design and framing of this research.

NL, RG, and MS were partially supported by cooperative agreement CDC-RFA-FT-23-0069 from the CDCs Center for Forecasting and Outbreak Analytics. AN was supported by the CDC-CFA award for the Atlantic Coast Center for Infectious Disease Dynamics and Analytics (ACCIDDA): NU38FT000012-01-00 (UNC). The views expressed in written in this publication do not necessarily reflect the official policies of the Department of Health and Human Services/Centers for Disease Control and Prevention.

## Author Contributions

N.B.L. designed the study, developed the analytical framework, wrote the code, and drafted the manuscript. M.S. supervised the work, contributed to funding acquisition, contributed to study design, and critically revised the manuscript. R.G. contributed to analytical framework, chart review and critically revised the manuscript. A.N. contributed to analytical framework and critically revised the manuscript. All authors approved the final version for submission.

## Notes

### Competing Interest Statement

The authors have declared no competing interest.

