## Supplementary Materials for "Wastewater Surveillance as an Event Detection System: Outbreak and Peak Detection of SARS-CoV-2 Across 281 U.S. Counties"

May 14, 2026

#### S1 Inter-rater agreement on curated case events

##### S1.1 Purpose and interpretation

We report inter-rater agreement for two related curation tasks on county-level outbreaks and peaks. Both tasks were reviewed independently by NBL (first rater) and RG (second rater) against algorithmic proposals, with edits recorded so that final outputs could be compared transparently.

NBL created the curated events for all locations and spatial scales in the main paper, while RG reviewed a subset of county-level events to assess the consistency of these labels.

For outbreaks, the primary comparison is between two final onset lists after curation, using detection-style metrics. To allow for some flexibility around the choice of start date for outbreaks, we allowed a three-week tolerance window (just like the three-week detection interval in the main results) that counted a match if reviewed outbreaks were within three weeks of each other.

Peaks results are summarized similarly to outbreaks. During review, peaks were only removed; none were added or had their date changed. This differs from outbreak review, where some events were added or had their date adjusted.

##### S1.2 Data sources and baseline caveat

NBL and RG each applied edits to algorithmically proposed outbreaks and peaks. For peaks, the algorithmic proposals used by NBL and RG were equivalent. However, for outbreaks the automatic proposals were not identical across workflows, because of small differences in case-series preprocessing and alignment (including slightly different effective date ranges at the ends of the series). A brief diagnostic on the pre-review algorithmic onset lists indicated that only a small fraction of automatically proposed events were affected at week-scale agreement (∼1%); hence, this misalignment has limited impact on the results.

##### S1.3 Results

**Outbreaks.** Table S1 compares final outbreak lists using a three-week matching window within county.

Table S1: Agreement between final outbreak lists (three-week window within county). TP = paired outbreaks; NBL-only = outbreak defined by NBL but not RG; RG-only = outbreak defined by RG but not NBL.

| Quantity | Symbol / definition | Value |
|---|---|---|
| True positives (pairs) | TP | 66 |
| NBL-only | – | 9 |
| RG-only | – | 16 |
| NBL events | $\lvert NBL \rvert$ | 75 |
| RG events | $\lvert RG \rvert$ | 82 |
| NBL match rate | TP/$\lvert NBL \rvert$ | 0.880 |
| RG match rate | TP/$\lvert RG \rvert$ | 0.805 |

Agreement is strong overall: 88% of NBL outbreaks were matched to an RG outbreak, and 80% of RG outbreaks were matched to an NBL outbreak.

**Peaks.** Table S2 compares final peak lists within county.

Table S2: Agreement between final peak lists (within county). TP = paired outbreaks; NBL-only = outbreak defined by NBL but not RG; RG-only = outbreak defined by RG but not NBL.

| Quantity | Symbol / definition | Value |
|---|---|---|
| True positives (pairs) | TP | 72 |
| NBL-only | – | 12 |
| RG-only | – | 10 |
| NBL events | $\lvert NBL \rvert$ | 84 |
| RG events | $\lvert RG \rvert$ | 82 |
| NBL match rate | TP/$\lvert NBL \rvert$ | 0.857 |
| RG match rate | TP/$\lvert RG \rvert$ | 0.878 |

Agreement is strong: 86% of NBL peaks were matched to an RG peak, and 88% of RG peaks were matched to an NBL peak.

##### S1.4 Summary

Both outbreak and peak comparisons indicate high inter-rater reliability under the stated tolerances, with residual discrepancy concentrated in a minority of events. At the same time, the remaining disagreements reflect the inherent subjectivity in defining outbreaks and peaks: even experienced reviewers will draw boundaries differently on ambiguous cases. This has a direct implication for algorithmic detection — the limiting factor is not model choice or implementation, but the absence of a fully agreed-upon ground truth.

#### S2 Model Convergence

We assessed model convergence using the Gelman-Rubin R-hat statistic [1] and the effective sample size (ESS), as shown in Figures S1 and S2. The R-hat statistic estimates the proportion of total to within-chain variance; values close to 1 indicate proper convergence. The ESS approximates the equivalent number of independent samples from the posterior that the MCMC fit achieves. With 900 total posterior samples, ESS values close to 900 indicate low autocorrelation and efficient sampling.

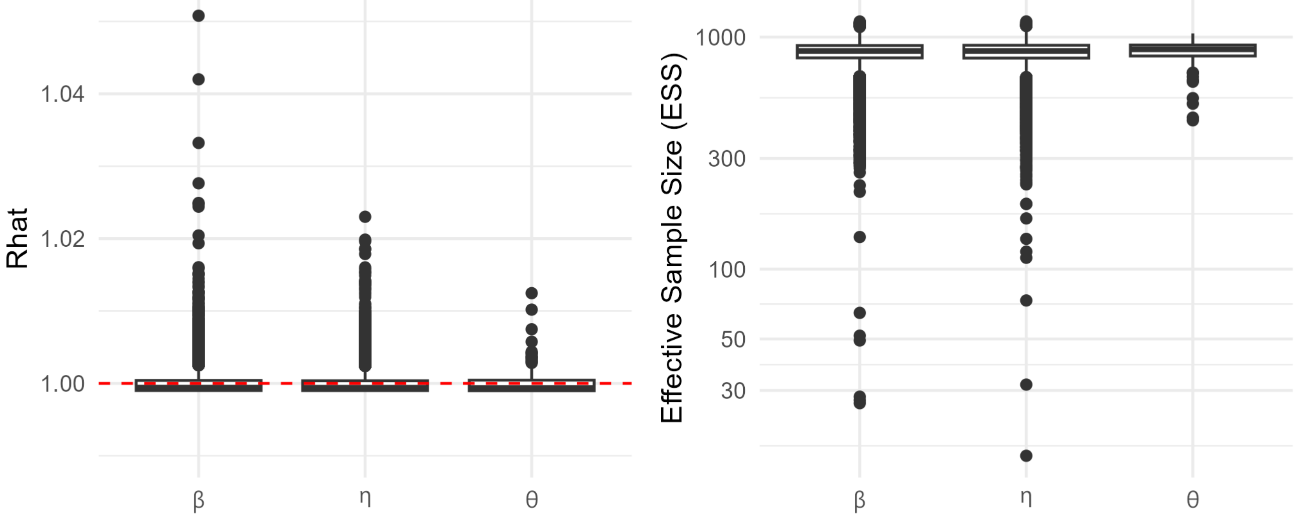


Figure S1: Boxplots of Gelman-Rubin R-hat statistic and effective sample sizes for the WW model runs across locations. For $\beta$ and $\eta$, each data point in the boxplot corresponds to a specific location and time (e.g. $\beta_{i,t}$), and for $\theta$ each point corresponds to a location-specific value ($\theta_i$). Boxes represent the interquartile range (IQR), with the median shown as a line inside the box; whiskers extend to the most extreme data points within 1.5 times the IQR from the first and third quartiles.

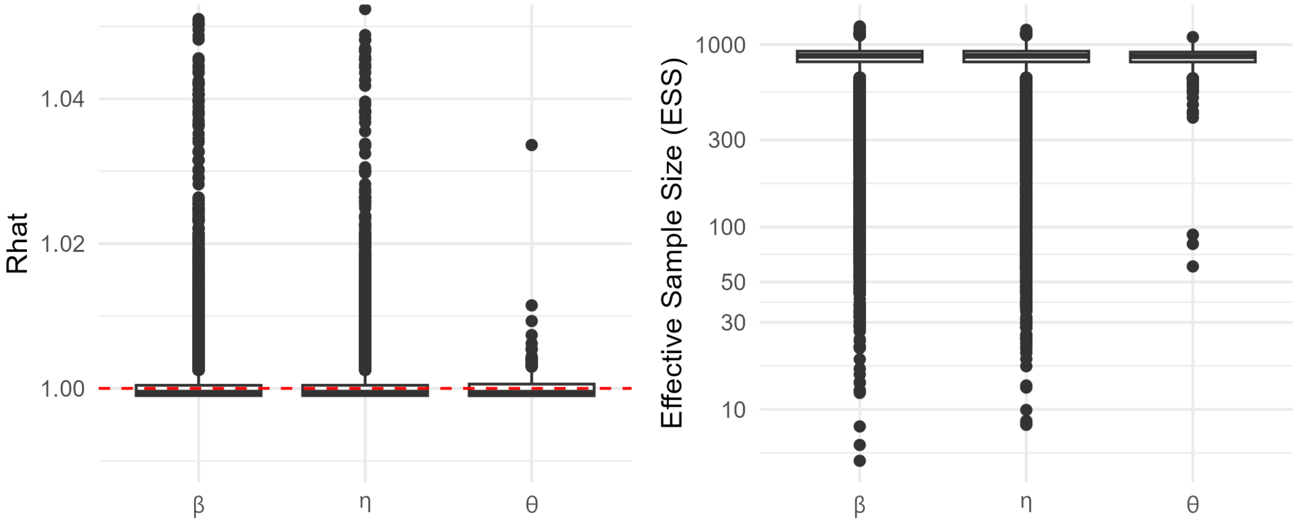


Figure S2: Boxplots of Gelman-Rubin R-hat statistic and effective sample sizes for the cases model runs across locations. For $\beta$ and $\eta$, each data point in the boxplot corresponds to a specific location and time (e.g. $\beta_{i,t}$), and for $\theta$ each point corresponds to a location-specific value ($\theta_i$). Boxes represent the interquartile range (IQR), with the median shown as a line inside the box; whiskers extend to the most extreme data points within 1.5 times the IQR from the first and third quartiles.

For the WW model, 99.7%, 99.8%, and 99.3% of R-hat values for $\beta$, $\eta$, and $\theta$ were below 1.01, indicating strong convergence across parameters. Additionally, the ESS exceeded 50 in over 99.9% of instances for all three parameters, further supporting adequate posterior mixing. For the cases model fit, 99.3% of R-hat values for $\beta$, $\eta$, and $\theta$ were below 1.01, indicating strong convergence. In addition, the ESS exceeded 50 in over 99.8% of instances for all three parameters.

In addition to R-hat and ESS, we assessed the number of divergent transitions in each model fit. Divergent transitions occur when the Hamiltonian trajectory deviates from the true posterior trajectory, indicating issues with posterior geometry or model specification [2]. Table S3 summarizes the number of model locations with any divergences, the average number of divergences, and the average divergences among locations with them.

The number of locations with divergences in the WW model was relatively high—45 in total. This can be largely attributed to missing data fed into the model. Among locations with complete data after interpolation, only 2.5% had any divergent transitions. In contrast, 92.9% of locations with gaps in the data experienced divergences. This likely reflects the model's difficulty estimating the relevant $\beta$ and $\eta$ parameters in windows with missing observations. While the presence of divergences is a concern, the model nonetheless appears to provide reasonable fits even in affected locations. This is reinforced by the high R-hat and ESS values for the parameters, even in locations with divergences.

The cases model had a similar number of divergences—47 in total. This is partially explained by data with zero values. Among the locations that had weeks with no cases reported, 51% had divergent transitions, while of the locations without any zero-valued weeks, there were 10.2% divergences. The model struggles to fit zero-valued observations, possibly because the negative binomial likelihood is sharp (i.e. highly penalized for mean estimates that slightly deviate from 0). As with the WW model, while divergent transitions are concerning, the model fits still appear reasonable for the case data, and the R-hat and ESS values indicate strong convergence.

| **Outcome** | **Locations** | # divergent locations (%) | Avg % of divergences | Avg % for locs with divergences |
|---|---|---|---|---|
| WW | With complete data | 6/239 (2.5%) | 0.1% | 4.4% |
| | With gaps | 39/42 (92.9%) | 4% | 4.3% |
| | Total | 45/281 (16%) | 0.7% | 4.3% |
| Cases | Without zero values | 24/236 (10.2%) | 0.5% | 5% |
| | With zero values | 23/45 (51.1%) | 3.7% | 7.2% |
| | Total | 47/281 (16.7%) | 1% | 6.1% |

Table S3: Summary of divergences for WW and cases. The divergences for WW locations with complete data after interpolation and with gaps are shown along with the total. For cases, the divergences for locations with and without any zero-valued counts in the data are shown along with the total. The columns show the total number of locations with divergences, the average number of divergences, and the average number of divergences per location that had any.

### S3 Outbreak Detection Comparison to $R_t$ Baseline

This section describes the $R_t$-based outbreak detection method used as a baseline comparison for the onset detection approach, and presents results across all method pairings.

#### S3.1 $R_t$ Estimation Comparison Model

As a baseline for comparison, we implement an outbreak detection method based on the effective reproductive number ($R_t$) that we call *deconv-Rt*. We estimate $R_t$ for wastewater data ($R_{t,ww}$) and reported case data ($R_{t,c}$). $R_t$ represents the average number of secondary infections generated by a primary infection, with $R_t > 1$ indicating epidemic growth and $R_t < 1$ indicating decline. $R_t$ is a standard metric used by researchers and public health officials to monitor epidemic trends and detect changes in disease transmission, making it a useful benchmark for practical event detection.

##### S3.1.1 Wastewater $R_{t,ww}$ Estimation

Using the methods from Huisman et al. [3, 4], we estimate $R_{t,ww}$ from wastewater data in a two-step process. First, wastewater measurements are deconvolved into estimated incidence curves using an expectation-maximization algorithm from Goldstein et al. [5]. This process estimates the most likely incidence curves that could have generated the wastewater time series. To capture uncertainty, the deconvolution step is bootstrapped to produce $n_{boot}$ incidence curve estimates. Second, for each incidence curve estimate, we apply the EpiEstim package [6] to estimate $R_{t,ww}$. While this method is optimally implemented for data with multiple samples per week, we use it for our weekly dataset given the absence of a better-validated alternative for this sampling frequency. This method requires as input a shedding load distribution (the profile of viral RNA shedding into wastewater from time of infection) for deconvolution and a serial interval distribution (time between successive cases) for $R_t$ estimation. We adopt the default parameters in [3] for both distributions, which are based on prior COVID-19 research. We set $n_{boot} = 200$.

From the $j = 1, \ldots, 200$ bootstrap iterations, we compute the following summaries:

- $\overline{R_{t,ww}} = \text{median}\left(\overline{R_{t,ww}(j)}\right)$
- $R^{0.25}_{t,ww} = \min\left(\text{median}_j\left(R^{0.25}_{t,ww}(j)\right),\ \text{median}_j\left(\overline{R_{t,ww}(j)}\right) - 0.675 \times \text{sd}_j\left(\overline{R_{t,ww}(j)}\right)\right)$
- $R^{0.05}_{t,ww} = \min\left(\text{median}_j\left(R^{0.05}_{t,ww}(j)\right),\ \text{median}_j\left(\overline{R_{t,ww}(j)}\right) - 1.645 \times \text{sd}_j\left(\overline{R_{t,ww}(j)}\right)\right)$

Here, $\overline{R_{t,ww}(j)}$ denotes the mean posterior $R_{t,ww}$ from EpiEstim in the $j^{\text{th}}$ bootstrap sample. Thus, $R^{0.05}_{t,ww}$ reflects both the median credible quantiles and quantiles of the bootstrapped means.

*deconv-Rt* produces daily estimates of $R_{t,ww}$. Outbreaks are defined analogously to the exponential growth model:

- *Option A:* An outbreak begins at $t$ if $R^{0.05}_{t,ww} > 1$ and no outbreak is ongoing.
- *Option B:* An outbreak begins at $t$ if $R^{0.25}_{t,ww} > 1$ and no outbreak is ongoing.
- An outbreak ends at $t$ if $\overline{R_{t,ww}} < 1$ while an outbreak is active.

We assess outbreak detection performance using both $R^{0.05}_{t,ww}$ and $R^{0.25}_{t,ww}$ thresholds, calling these *deconv-Rt(0.05)* and *deconv-Rt(0.25)* respectively. Although $R^{0.05}_{t,ww}$ matches the quantile used for the exponential growth model, it detects few outbreaks and performs poorly. $R^{0.25}_{t,ww}$ yields more detections and better performance. Because EpiEstim only returns 0.025, 0.05, and 0.25 lower quantiles, no additional quantile thresholds were evaluated.

##### S3.1.2 Cases $R_{t,c}$ Estimation

The $R_{t,c}$ estimates for cases were generated using methods analogous to those applied to wastewater data, following the approach of Huisman et al. [4, 3]. Instead of the shedding load distribution used for wastewater, cases were deconvolved into estimated incidence curves using the distribution of the incubation period (delay from infection to symptoms) and the distribution of time from symptoms to clinical case reporting. We used the default incubation and reporting delay distributions provided by Huisman et al. [3]. All remaining steps in the $R_{t,c}$ estimation process were identical to those used for wastewater data.

*deconv-Rt(0.05)* and *deconv-Rt(0.25)* are fit on wastewater and case data for all counties in our study. We assess multiple wastewater–cases method pairings, classifying the detected outbreaks according to the rules described in Section 3.4 of the main text.

#### S3.2 Results

Tables S4 and S5 show outbreak detection performance across all 281 counties and the subset of 184 counties with populations of at least 100,000, respectively, for all wastewater–cases method pairings. The *exp growth–exp growth* pairing achieves the strongest performance (sensitivity 0.82, PPV 0.64 across all counties), substantially outperforming all *deconv-Rt* variants. Replacing *exp growth* with *deconv-Rt(0.25)* in either data stream reduces both sensitivity and PPV, likely due to a combination of worse detection and misaligned outbreak timing between methods. Misaligned timing can occur because *deconv-Rt(0.25)* may detect outbreaks earlier than *exp growth* owing to its daily $R_t$ estimates, as evidenced by the negative mean lead time when using *exp growth* on wastewater and *deconv-Rt(0.25)* on cases. Importantly, even when using *deconv-Rt(0.25)* for both wastewater and cases, performance remains poor (PPV = 0.19), suggesting that *deconv-Rt* produces event series that are less well aligned between data streams and therefore less useful for reliable outbreak alerting in this weekly setting. Performance declines further with *deconv-Rt(0.05)*, which detects too few outbreaks to be useful, with near-zero sensitivity and PPV rendering it impractical for outbreak detection in this context.

| Wastewater Method | Cases Method | Detection Interval | Sensitivity | PPV | TPs | FPs | FNs | Mean Lead |
|---|---|---|---|---|---|---|---|---|
| *exp growth* | *exp growth* | $[-3, 3]$ | **0.82** | **0.64** | 306 | 175 | 67 | 0.62 |
| *exp growth* | *deconv-Rt(0.25)* | $[-3, 3]$ | 0.58 | 0.28 | 142 | 373 | 102 | -0.56 |
| *exp growth* | *deconv-Rt(0.05)* | $[-3, 3]$ | 0.44 | 0.02 | 11 | 517 | 14 | -0.83 |
| *deconv-Rt(0.25)* | *exp growth* | $[-3, 3]$ | 0.50 | 0.27 | 202 | 560 | 206 | 0.57 |
| *deconv-Rt(0.25)* | *deconv-Rt(0.25)* | $[-3, 3]$ | 0.58 | 0.19 | 161 | 672 | 115 | 0.83 |
| *deconv-Rt(0.05)* | *exp growth* | $[-3, 3]$ | 0.08 | 0.23 | 33 | 112 | 381 | 0.47 |
| *deconv-Rt(0.05)* | *deconv-Rt(0.05)* | $[-3, 3]$ | 0 | 0 | 0 | 154 | 27 | NA |

Table S4: **County-level** classification metrics across different wastewater and cases methods. *exp growth* refers to the exponential growth model introduced in this study; *deconv-Rt(0.25)* and *deconv-Rt(0.05)* refer to the $R_t$ estimation methods with outbreak detection cutoffs of 0.25 and 0.05 respectively. The detection interval is the allotted window in weeks where an indicator outbreak is treated as a true positive of the outcome. The columns show the sensitivity, positive predictive value (PPV), number of true positives (TPs), false positives (FPs), false negatives (FNs), and mean lead, defined as the average time the wastewater outbreak occurs before the cases outbreak (in weeks) for TPs. Positive values indicate the wastewater outbreak occurs first.

#### S3.3 Why Exponential Growth Outperforms the $R_t$ Baseline

Several structural differences between *exp growth* and *deconv-Rt* help explain the performance gap. While *exp growth* is a Bayesian model that assumes a particular likelihood and prior distributions, it does not require externally specified fixed parameters. In contrast, *deconv-Rt* relies on predefined inputs such as shedding load distributions, infection-to-case delays, and serial intervals—all of which introduce potential for misspecification. Additionally, *deconv-Rt* follows a multi-step process involving deconvolution and subsequent $R_t$ estimation, which introduces further uncertainty. The simplicity and single-stage structure of *exp growth* may be advantageous, especially with temporally sparse (i.e., weekly) data, where a more parsimonious model can reduce variability and risk of overfitting.

The posterior credible intervals produced by *exp growth* provide a coherent Bayesian measure of uncertainty. In contrast, the uncertainty intervals from *deconv-Rt* lack a clear statistical interpretation: they are formed by combining median credible intervals and bootstrapped medians, but do not correspond to standard confidence or credible intervals. This ambiguous uncertainty quantification may hinder accurate outbreak detection. Additionally, *deconv-Rt* only reports lower quantile values of 0.025, 0.05, and 0.25, which limits fine-tuning of the outbreak detection cutoff. *exp growth* models outcomes using a negative binomial distribution, which is more appropriate for overdispersed count data; by contrast, *deconv-Rt* via EpiEstim assumes a Poisson distribution, which is often too restrictive and can lead to poor model fit.

| Wastewater Method | Cases Method | Detection Interval | Sensitivity | PPV | TPs | FPs | FNs | Mean Lead |
|---|---|---|---|---|---|---|---|---|
| *exp growth* | *exp growth* | $[-3, 3]$ | 0.88 | **0.69** | 240 | 109 | 33 | 0.68 |
| *exp growth* | *curated cases* | $[-3, 3]$ | **0.92** | 0.57 | 196 | 147 | 17 | 0.72 |
| *exp growth* | *deconv-Rt(0.25)* | $[-3, 3]$ | 0.61 | 0.23 | 87 | 286 | 55 | -0.35 |
| *exp growth* | *deconv-Rt(0.05)* | $[-3, 3]$ | 0.37 | 0.02 | 7 | 375 | 12 | -0.45 |
| *deconv-Rt(0.25)* | *exp growth* | $[-3, 3]$ | 0.45 | 0.30 | 137 | 316 | 166 | 0.69 |
| *deconv-Rt(0.25)* | *curated cases* | $[-3, 3]$ | 0.46 | 0.23 | 103 | 354 | 122 | 0.82 |
| *deconv-Rt(0.25)* | *deconv-Rt(0.25)* | $[-3, 3]$ | 0.46 | 0.15 | 77 | 432 | 89 | 0.68 |
| *deconv-Rt(0.05)* | *exp growth* | $[-3, 3]$ | 0.06 | 0.33 | 18 | 37 | 290 | 0.76 |
| *deconv-Rt(0.05)* | *curated cases* | $[-3, 3]$ | 0.07 | 0.28 | 15 | 38 | 214 | 0.84 |
| *deconv-Rt(0.05)* | *deconv-Rt(0.05)* | $[-3, 3]$ | 0 | 0 | 0 | 59 | 20 | NA |

Table S5: **County-level** classification metrics across different wastewater and cases methods **for counties with population of at least 100,000**. *exp growth* refers to the exponential growth model introduced in this study, *curated cases* refers to the curated cases defined in Section 3.3 of the main text, and *deconv-Rt(0.25)* and *deconv-Rt(0.05)* refer to the $R_t$ estimation methods with outbreak detection cutoffs of 0.25 and 0.05 respectively. The detection interval is the allotted window in weeks where an indicator outbreak is treated as a true positive of the outcome. The columns show the sensitivity, positive predictive value (PPV), number of true positives (TPs), false positives (FPs), false negatives (FNs), and mean lead, defined as the average time the wastewater outbreak occurs before the cases outbreak (in weeks) for TPs. Positive values indicate the wastewater outbreak occurs first.

Another limitation of *deconv-Rt* is that it estimates daily $R_t$ values and may not be well suited to the weekly data used in this study. Prior work has shown that its performance deteriorates with fewer than three wastewater samples per week [3], making its outbreak detection less reliable and more unstable in such settings. Furthermore, since *deconv-Rt* operates on a shorter time scale, it may detect outbreaks earlier than *exp growth*, which uses a five-week fitting window. However, early detection alone does not account for the inferior performance of *deconv-Rt*, as the wastewater-cases pairing using *deconv-Rt*—which shares the same time scale—still performs poorly. Overall, *exp growth* appears to be a more robust and practical method for outbreak detection using weekly wastewater data.

#### S4 Peak Detection Method Comparisons

We evaluated a range of peak detection configurations, including different window sizes and three classes of signal-based filters: a peakiness-noisiness score, a quantile threshold, and a piecewise slope test. Results are presented across all counties (against algorithmically-defined case peaks) and for counties with populations of at least 100,000 (against curated case peaks).

Table ?? presents results across window sizes ranging from $w = 3$ to $w = 8$. Performance declines monotonically as the window grows: the $w = 3$ symmetric window achieves the highest sensitivity (0.83) and PPV (0.69) across all counties, and similarly leads on sensitivity (0.87) against curated peaks in larger counties (Table ??), though PPV is lower in that setting due to the higher density of wastewater peaks relative to curated case peaks. These results suggest that a simple peak detection approach—taking the maximum value within a symmetric window of 3 weeks on each side of the candidate point—is an effective and robust baseline.

| Peak detection method | Sensitivity | PPV | TP | FP | FN |
|---|---|---|---|---|---|
| w = 3 | **0.83** | **0.70** | 572 | 249 | 114 |
| w = 4 | 0.79 | 0.67 | 431 | 213 | 114 |
| w = (5, 3) | 0.81 | 0.65 | 426 | 233 | 103 |
| w = 5 | 0.74 | 0.63 | 328 | 195 | 114 |
| w = 6 | 0.73 | 0.62 | 287 | 174 | 105 |
| w = 7 | 0.70 | 0.62 | 252 | 152 | 108 |
| w = (8, 3) | 0.78 | 0.61 | 320 | 202 | 91 |
| w = 8 | 0.67 | 0.62 | 219 | 137 | 110 |

Table S6: Peak detection window size comparison. The peak detection approach is applied to both wastewater and cases data; true positives (TP) are classified when peaks occur three weeks or less apart. Methods are named as $w = k$ for symmetric windows or $w = (k_L, k_R)$ for asymmetric windows, where $k_L$ is the left window and $k_R$ is the right window in weeks. All window sizes are evaluated on the same dataset by censoring based on $w = 8$, meaning peak detection is only performed on data points $[9, n - 8]$ where $n$ is the total number of observations.

| Peak detection method | Sensitivity | PPV | TP | FP | FN |
|---|---|---|---|---|---|
| w = 3 (censored with quantile 10) | **0.89** | 0.42 | 274 | 379 | 33 |
| w = 3 with quantile(10, 0.5) | 0.89 | 0.45 | 271 | 337 | 35 |
| w = 3 with quantile(10, 0.7) | 0.85 | 0.46 | 261 | 302 | 45 |
| w = 3 with quantile(10, 0.8) | 0.85 | 0.48 | 255 | 275 | 46 |
| w = 3 with quantile(10, 0.9) | 0.83 | **0.51** | 244 | 238 | 51 |
| w = 3 (censored with quantile 15) | **0.89** | 0.42 | 232 | 324 | 28 |
| w = 3 with quantile(15, 0.5) | 0.88 | 0.45 | 230 | 276 | 30 |
| w = 3 with quantile(15, 0.7) | 0.88 | 0.48 | 225 | 246 | 32 |
| w = 3 with quantile(15, 0.8) | 0.86 | 0.50 | 219 | 222 | 37 |
| w = 3 with quantile(15, 0.9) | 0.84 | **0.52** | 209 | 191 | 41 |
| w = 3 (censored with quantile 20) | **0.90** | 0.42 | 200 | 272 | 21 |
| w = 3 with quantile(20, 0.5) | 0.90 | 0.45 | 198 | 239 | 23 |
| w = 3 with quantile(20, 0.7) | 0.88 | 0.48 | 194 | 210 | 26 |
| w = 3 with quantile(20, 0.8) | 0.84 | 0.49 | 184 | 189 | 34 |
| w = 3 with quantile(20, 0.9) | 0.80 | **0.51** | 170 | 161 | 42 |

Table S7: Peak detection with quantile filtering using curated cases peaks for counties with population at least 100,000. The peak detection approach is applied to wastewater data and true positives (TP) are classified against curated case peak dates when peaks occur three weeks or less apart. Methods are named as $w = k$ with quantile($l$, $q$), where $w$ is the window size in weeks, $l$ is the lookback period in weeks used to compute quantiles, and $q$ is the quantile threshold (a peak must exceed the $q$-th quantile of the lookback period to be counted). The baseline rows show $w = k$ (censored with quantile $l$), which applies the same censoring as quantile filtering.

We then evaluated three classes of filters applied on top of the $w = 3$ baseline to assess whether additional signal processing could improve detection. None improved on the unfiltered baseline.

**Peakiness–noisiness score.** For a candidate peak at time $t_p$ (index $i$), let $W$ denote the vector of values $y$ on the index window $[i - w_L,\, i + w_R]$, *including* $y(t_p)$, after dropping missing values. Let $W_{\text{left}}$ and $W_{\text{right}}$ denote the values on $[i - w_L,\, i]$ and $[i,\, i + w_R]$, respectively (each including the peak). Write $\Delta W_{\text{left}}$ and $\Delta W_{\text{right}}$ for the first-difference sequences along those segments, and range($W_{\text{left}}$), range($W_{\text{right}}$) for max − min when that range is positive (otherwise the corresponding noise term is set to zero in the implementation). We defined

$$\text{score}(t_p) = \lambda \cdot \frac{y(t_p) - \overline{W}}{\text{sd}(W)} - \frac{w_L}{w_L + w_R} \cdot \frac{\text{sd}(\Delta W_{\text{left}})}{\text{range}(W_{\text{left}})} - \frac{w_R}{w_L + w_R} \cdot \frac{\text{sd}(\Delta W_{\text{right}})}{\text{range}(W_{\text{right}})}, \tag{1}$$

where the noise ratios are omitted (treated as zero) if a segment has fewer than two points, fewer than two first differences, or zero range. The first term rewards prominence of $y(t_p)$ relative to the local mean and variability; the subtracted terms penalize choppiness on each side of the peak, combined with weights proportional to $w_L$ and $w_R$ (for a symmetric window $w_L = w_R = w$, each weight is 1/2). The parameter $\lambda$ scales prominence relative to this noise penalty. Candidates were retained when score($t_p$) exceeded a chosen threshold $\tau$ (in addition to the baseline local-maximum rules).[1]

Despite a grid search over $\lambda$ and the score threshold, and evaluation both with and without applying the same filter to case data, this approach did not improve peak detection performance (Tables S8 and S9).

**Piecewise slope filter.** For each candidate peak at index $i$, we fit a piecewise linear model over the window $[i - w_L, i + w_R]$ to test for a significant change in slope at the peak:

$$y_j = \beta_0 + \beta_1(j - i) + \beta_2(j - i) \cdot \mathbf{1}_{j>i} + \epsilon_j, \tag{2}$$

where $j \in [i - w_L, i + w_R]$ and $\mathbf{1}_{j>i}$ is an indicator for observations after the candidate peak. The interaction term $\beta_2$ captures the change in slope at the peak. Candidate peaks were retained when $\min(p_{\text{linear}}, p_{\text{log}}) < \alpha$, where the log model uses $\log(y_j)$ as the response. Despite its theoretical appeal as a test for trend reversal, this filter substantially reduced sensitivity without commensurate gains in PPV across all significance levels and test types evaluated (Tables S8 and S9).

| Peak detection method | Sensitivity | PPV | TP | FP | FN |
|---|---|---|---|---|---|
| w = 3 | **0.84** | **0.70** | 779 | 338 | 151 |
| w = 3 with score(0.1, -0.5) | 0.73 | 0.62 | 552 | 337 | 207 |
| w = 3 with score(0.1, -0.25) | 0.37 | 0.32 | 108 | 229 | 187 |
| w = 3 with score(0.2, -0.5) | 0.81 | 0.67 | 701 | 348 | 166 |
| w = 3 with score(0.2, -0.25) | 0.64 | 0.50 | 363 | 363 | 202 |
| w = 3 with score(0.3, -0.5) | 0.83 | 0.69 | 757 | 340 | 157 |
| w = 3 with score(0.3, -0.25) | 0.75 | 0.62 | 574 | 354 | 191 |
| w = 3 with slope(0.05, linear) | 0.37 | 0.45 | 132 | 161 | 227 |
| w = 3 with slope(0.1, linear) | 0.48 | 0.51 | 229 | 222 | 245 |
| w = 3 with slope(0.2, linear) | 0.61 | 0.58 | 383 | 274 | 244 |
| w = 3 with slope(0.05, log) | 0.38 | 0.44 | 143 | 181 | 233 |
| w = 3 with slope(0.1, log) | 0.45 | 0.50 | 221 | 223 | 266 |
| w = 3 with slope(0.2, log) | 0.58 | 0.57 | 351 | 265 | 253 |
| w = 3 with slope(0.05, both) | 0.40 | 0.46 | 156 | 184 | 231 |
| w = 3 with slope(0.1, both) | 0.48 | 0.50 | 240 | 239 | 255 |
| w = 3 with slope(0.2, both) | 0.62 | 0.58 | 391 | 286 | 242 |

Table S8: Peak detection with score and slope filtering. The peak detection approach is applied to both wastewater and cases data; true positives (TP) are classified when peaks occur three weeks or less apart. Methods are named as $w = k$ with filter($p_1$, $p_2$), where $w$ is the window size in weeks. For score filtering, $p_1$ is the smoothing parameter $\lambda$ and $p_2$ is the score threshold. For slope filtering, $p_1$ is the significance level $\alpha$ and $p_2$ is the test type (linear, log, or both).

**Quantile threshold filter.** A candidate peak was retained only if its value exceeded a specified quantile (e.g., the 80th percentile) of wastewater measurements over a preceding lookback window of $N$ weeks. Across all lookback lengths and quantile thresholds evaluated, this filter did not improve on the unfiltered baseline (Tables S10 and S11).

[1] Mean and SD may instead be computed on $W \setminus \{y(t_p)\}$ if the peak is excluded from the window; the default follows inclusion.

| Peak detection method | Sensitivity | PPV | TP | FP | FN |
|---|---|---|---|---|---|
| w = 3 | **0.88** | 0.40 | 307 | 466 | 40 |
| w = 3 with score(0.1, -0.5) | 0.80 | 0.43 | 270 | 354 | 69 |
| w = 3 with score(0.1, -0.25) | 0.41 | 0.53 | 131 | 116 | 187 |
| w = 3 with score(0.2, -0.5) | 0.86 | 0.41 | 297 | 428 | 48 |
| w = 3 with score(0.2, -0.25) | 0.69 | 0.44 | 226 | 283 | 103 |
| w = 3 with score(0.3, -0.5) | 0.88 | 0.40 | 304 | 455 | 43 |
| w = 3 with score(0.3, -0.25) | 0.80 | 0.43 | 273 | 368 | 67 |
| w = 3 with slope(0.05, linear) | 0.40 | **0.56** | 126 | 100 | 187 |
| w = 3 with slope(0.1, linear) | 0.55 | 0.53 | 174 | 156 | 145 |
| w = 3 with slope(0.2, linear) | 0.69 | 0.49 | 228 | 241 | 103 |
| w = 3 with slope(0.05, log) | 0.43 | 0.55 | 134 | 111 | 181 |
| w = 3 with slope(0.1, log) | 0.53 | 0.52 | 169 | 156 | 151 |
| w = 3 with slope(0.2, log) | 0.67 | 0.49 | 218 | 230 | 109 |
| w = 3 with slope(0.05, both) | 0.44 | 0.54 | 140 | 117 | 175 |
| w = 3 with slope(0.1, both) | 0.56 | 0.52 | 180 | 167 | 140 |
| w = 3 with slope(0.2, both) | 0.70 | 0.48 | 232 | 254 | 99 |

Table S9: Peak detection with score and slope filtering using curated cases peaks for counties with population at least 100,000. The peak detection approach is applied to wastewater data and true positives (TP) are classified against curated case peak dates when peaks occur three weeks or less apart. Methods are named as $w = k$ with filter($p_1$, $p_2$), where $w$ is the window size in weeks. For score filtering, $p_1$ is the smoothing parameter $\lambda$ and $p_2$ is the score threshold. For slope filtering, $p_1$ is the significance level $\alpha$ and $p_2$ is the test type (linear, log, or both).

| Peak detection method | Sensitivity | PPV | TP | FP | FN |
|---|---|---|---|---|---|
| w = 3 (censored with quantile 10) | 0.83 | **0.71** | 652 | 263 | 130 |
| w = 3 with quantile(10, 0.5) | **0.84** | 0.68 | 582 | 271 | 111 |
| w = 3 with quantile(10, 0.7) | 0.83 | 0.66 | 518 | 264 | 106 |
| w = 3 with quantile(10, 0.8) | 0.83 | 0.66 | 485 | 253 | 101 |
| w = 3 with quantile(10, 0.9) | 0.81 | 0.64 | 426 | 240 | 101 |
| w = 3 (censored with quantile 15) | 0.84 | **0.72** | 555 | 217 | 103 |
| w = 3 with quantile(15, 0.5) | **0.86** | 0.70 | 489 | 212 | 79 |
| w = 3 with quantile(15, 0.7) | 0.85 | 0.68 | 442 | 207 | 79 |
| w = 3 with quantile(15, 0.8) | 0.84 | 0.64 | 386 | 219 | 73 |
| w = 3 with quantile(15, 0.9) | 0.83 | 0.59 | 323 | 220 | 68 |
| w = 3 (censored with quantile 20) | 0.85 | **0.73** | 460 | 170 | 83 |
| w = 3 with quantile(20, 0.5) | **0.86** | 0.69 | 405 | 178 | 66 |
| w = 3 with quantile(20, 0.7) | 0.85 | 0.60 | 321 | 214 | 57 |
| w = 3 with quantile(20, 0.8) | 0.82 | 0.50 | 247 | 247 | 53 |
| w = 3 with quantile(20, 0.9) | 0.81 | 0.40 | 171 | 260 | 41 |

Table S10: Peak detection with quantile filtering. The peak detection approach is applied to both wastewater and cases data; true positives (TP) are classified when peaks occur three weeks or less apart. Methods are named as $w = k$ with quantile($l$, $q$), where $w$ is the window size in weeks, $l$ is the lookback period in weeks used to compute quantiles, and $q$ is the quantile threshold (a peak must exceed the $q$-th quantile of the lookback period to be counted). The baseline rows show $w = k$ (censored with quantile $l$), which applies the same censoring as quantile filtering.

| Peak detection method | Sensitivity | PPV | TP | FP | FN |
|---|---|---|---|---|---|
| w = 3 (censored with quantile 10) | **0.89** | 0.42 | 274 | 379 | 33 |
| w = 3 with quantile(10, 0.5) | 0.89 | 0.45 | 271 | 337 | 35 |
| w = 3 with quantile(10, 0.7) | 0.85 | 0.46 | 261 | 302 | 45 |
| w = 3 with quantile(10, 0.8) | 0.85 | 0.48 | 255 | 275 | 46 |
| w = 3 with quantile(10, 0.9) | 0.83 | **0.51** | 244 | 238 | 51 |
| w = 3 (censored with quantile 15) | **0.89** | 0.42 | 232 | 324 | 28 |
| w = 3 with quantile(15, 0.5) | 0.88 | 0.45 | 230 | 276 | 30 |
| w = 3 with quantile(15, 0.7) | 0.88 | 0.48 | 225 | 246 | 32 |
| w = 3 with quantile(15, 0.8) | 0.86 | 0.50 | 219 | 222 | 37 |
| w = 3 with quantile(15, 0.9) | 0.84 | **0.52** | 209 | 191 | 41 |
| w = 3 (censored with quantile 20) | **0.90** | 0.42 | 200 | 272 | 21 |
| w = 3 with quantile(20, 0.5) | 0.90 | 0.45 | 198 | 239 | 23 |
| w = 3 with quantile(20, 0.7) | 0.88 | 0.48 | 194 | 210 | 26 |
| w = 3 with quantile(20, 0.8) | 0.84 | 0.49 | 184 | 189 | 34 |
| w = 3 with quantile(20, 0.9) | 0.80 | **0.51** | 170 | 161 | 42 |

Table S11: Peak detection with quantile filtering using curated cases peaks for counties with population at least 100,000. The peak detection approach is applied to wastewater data and true positives (TP) are classified against curated case peak dates when peaks occur three weeks or less apart. Methods are named as $w = k$ with quantile($l$, $q$), where $w$ is the window size in weeks, $l$ is the lookback period in weeks used to compute quantiles, and $q$ is the quantile threshold (a peak must exceed the $q$-th quantile of the lookback period to be counted). The baseline rows show $w = k$ (censored with quantile $l$), which applies the same censoring as quantile filtering.

#### S5 Lead Time Distributions for County-Level Outbreaks and Peaks

The histograms in Figures S3 and S4 show the distribution of lead times for true positive outbreak and peak detections, respectively, at the county-level, providing a more granular view of the timing relationships summarized by the mean lead times reported in the main text.

The detection interval for case-based event detection was $[-3, 3]$; however, the lead time (or lag) between wastewater and cases varies within this interval. Figure S3 shows the distribution of lead times for outbreak TPs, where a positive lead time indicates wastewater events preceding cases events. The distribution is nearly identical for model-defined case outbreaks and manually-defined ones, with wastewater outbreaks typically leading or synchronous with cases. Most lead times are between 0 and 2 weeks, consistent with the expected population-level lag between viral shedding in wastewater and case detection and reporting.

The peak lead times show a less clear pattern. For the algorithmic peak comparison, a 1-week lead time is still the most frequent, but lead and lag times are broadly spread across $-3$ to 3 weeks. There is a clearer pattern in the curated case peaks plot, with lead times more concentrated around 1 week.

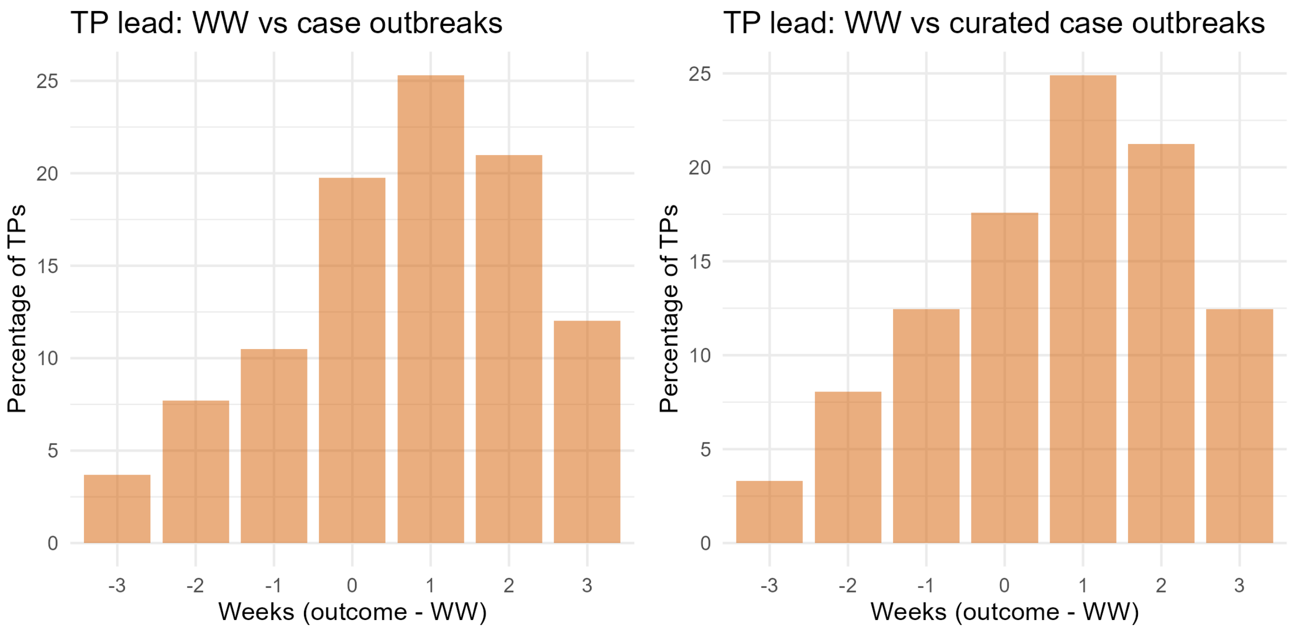


Figure S3: County-level TP lead-time distribution between wastewater (WW) and case outbreaks. The histogram shows the week offset $t$ = (case outbreak/outcome date) $-$ (WW date) in integer weeks, binned to $t \in [-3, 3]$ and plotted as the percentage of true positives (TPs); positive values indicate that WW precedes the case outbreak. The left/right panels use onset-detection-estimated case outbreaks vs curated case outbreaks, respectively.

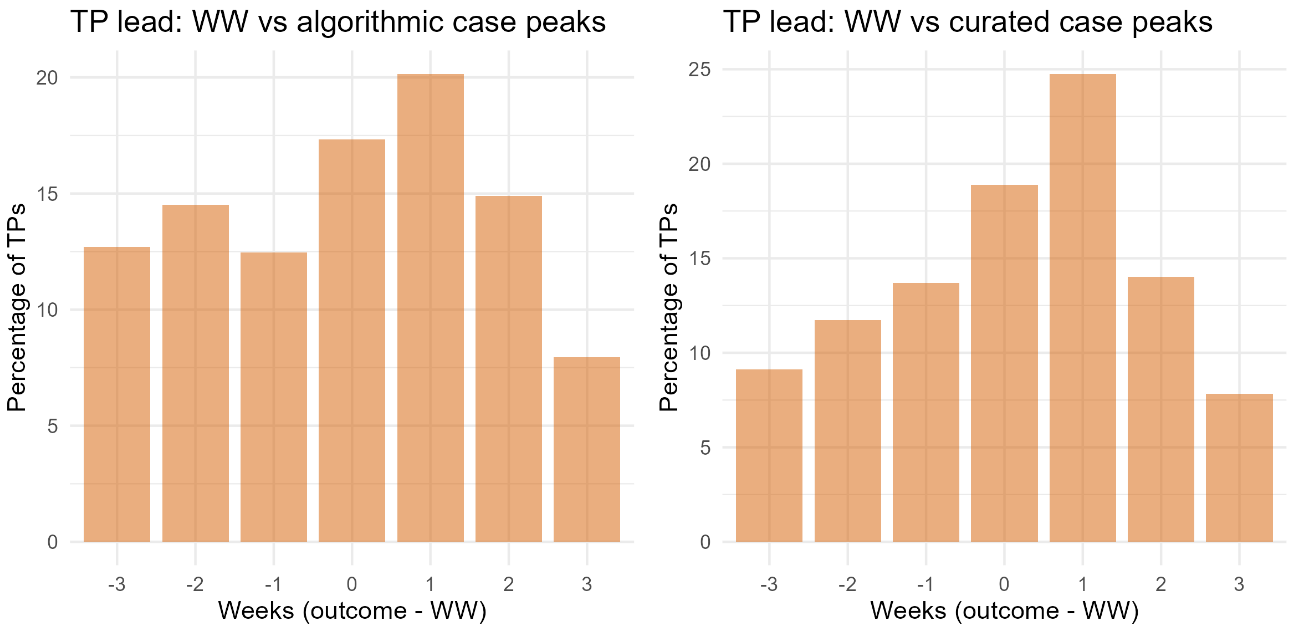


Figure S4: County-level TP lead-time distribution between wastewater (WW) and case peak dates. The histogram shows the week offset $t$ = (case peak date) $-$ (WW peak date) in integer weeks, binned to $t \in [-3, 3]$ and plotted as the percentage of true positives (TPs); positive values indicate that WW precedes the case peak. The left/right panels compare WW vs algorithmic case peaks and WW vs curated case peaks, respectively.

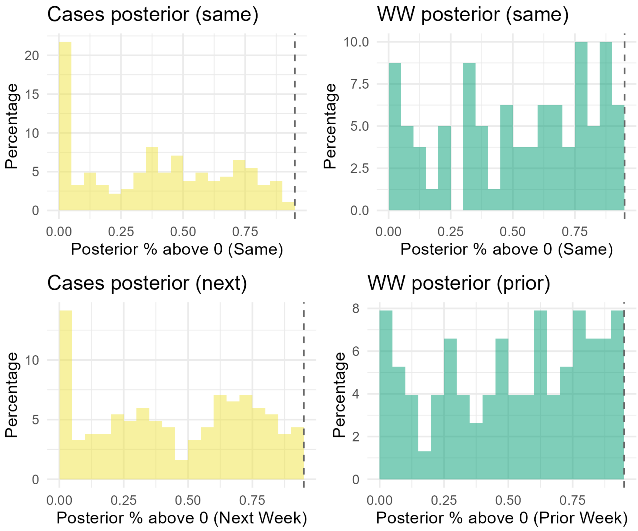


Figure S5: Distribution of the posterior probability that growth rate $\eta > 0$ (exponential growth model) in the opposing data stream at **onset detection** false positives and false negatives. Top row: for each false positive (FP), the cases-model posterior % above 0 in the same week (left); for each false negative (FN), the wastewater-model posterior % above 0 in the same week (right). Bottom row: for each FP, the cases-model posterior % above 0 in the next week (left); for each FN, the wastewater-model posterior % above 0 in the prior week (right). Histograms show the percentage of FP/FN events at each posterior % (0–0.95).

#### S6 Characterizing False Positives and False Negatives

To better understand the nature of misclassifications, we examine the characteristics of false positives (FPs) and false negatives (FNs) for both the onset detection and peak detection approaches. Rather than treating FPs and FNs as uniform errors, these analyses ask whether misclassified events show any systematic signal in the opposing data stream (e.g. cases relative to FPs and wastewater relative to FNs), suggesting they may reflect near-threshold events rather than outright detection failures.

Figure S5 examines FPs and FNs from the onset detection approach through the lens of the opposing data stream's posterior growth rate. For onset detection FPs—wastewater outbreaks with no corresponding case outbreak—the cases model shows a high concentration of posteriors falling entirely below zero, indicating strong misalignment between the two data streams in those instances, possibly reflecting overly confident model fitting. Among the remaining FPs, the distribution of the posterior probability of positive growth is broadly spread. Examining the cases model one week after the wastewater outbreak, the share of posteriors entirely below zero remains high, though more posteriors show moderate-to-high probability of positive growth. This suggests that some FPs represent wastewater signals that slightly precede a case-based increase that falls just short of the outbreak threshold, while others reflect genuine misalignment with case-based dynamics. For FNs—case outbreaks with no corresponding wastewater detection—the wastewater model's posterior probability of positive growth in the same week is broadly distributed, with a notable concentration of moderate-to-high values. The week prior shows a slightly more concentrated distribution of moderate-to-high posterior growth probabilities, suggesting that many FNs occur when wastewater is growing in the week preceding the case outbreak but does not reach the 95th percentile threshold required to declare an onset.

Figure S6 examines FPs and FNs from the peak detection approach by comparing the quantile of the opposing data stream at the time of the misclassification. Peak detection FPs—wastewater peaks with no matching case peak—tend to concentrate case quantiles around the middle point (the fourth largest out of seven points). Peak detection FNs—case peaks not matched by a wastewater peak—show a broadly distributed wastewater quantile in the same week, but many have high wastewater quantiles in the prior week. This indicates that many FNs are near-misses where wastewater almost reaches a peak in the preceeding week.

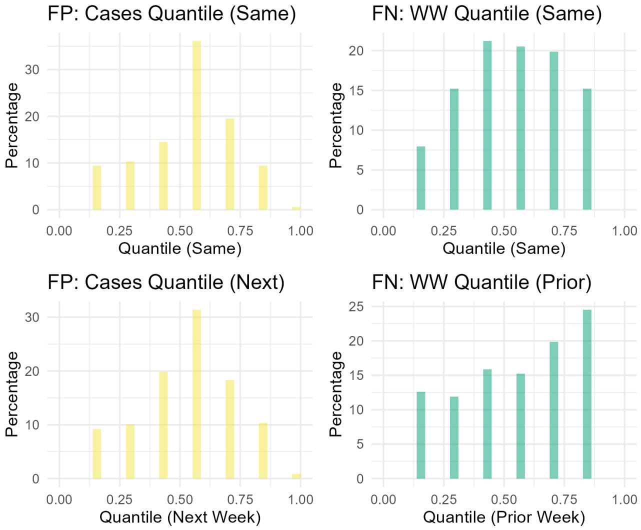


Figure S6: Distribution of case and wastewater quantiles at **peak detection** false positives and false negatives. Top row: for each false positive (FP), the quantile of cases in the same week (left); for each false negative (FN), the quantile of wastewater in the same week (right). Bottom row: for each FP, the quantile of cases in the next week (left); for each FN, the quantile of wastewater in the prior week (right). Histograms show the percentage of FP/FN events at each quantile (0–1). FP quantile reaches 1 only when the corresponding cases peak was excluded for having fewer than 10 cases.

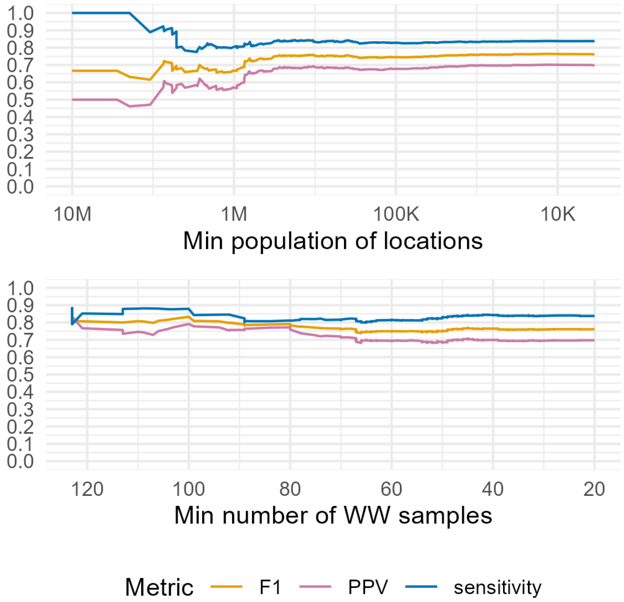


Figure S7: Rolling **peak detection** performance by location inclusion. Top: PPV, sensitivity, and F1 when locations are added in descending order of minimum population (x-axis, log scale; high population on the left). Bottom: same metrics when locations are added in descending order of minimum number of wastewater samples. Each point represents performance aggregated over all locations included up to that x-axis value.

### S7 Event Detection by Population Size and Wastewater Samples

Detection performance may vary systematically with location characteristics, such as population size and the number of wastewater samples collected. Larger counties tend to have greater signal-to-noise in both case reporting (and potentially with wastewater surveillance), while locations with more overall sampling may yield more stable event detection. To assess this, Figures S7 and S8 show rolling detection performance as locations are successively added in descending order of population and sample count.

For peak detection (Figure S7), performance is relatively stable across the full range of locations and wastewater sample counts, suggesting that the peak detection approach is broadly applicable regardless of county size.

For outbreak detection (Figure S8), there is a more pronounced trend: performance is stronger among higher-population counties and declines as smaller counties are included. This likely reflects two contributing factors. First, case reporting in larger counties may be more complete, yielding more accurate case-based outbreak definitions. Second, larger counties may have larger catchment areas that reduce noise in the wastewater signal. The latter is difficult to verify without detailed information on the sampling characteristics at each location, but the former seems plausible given known patterns in surveillance capacity.

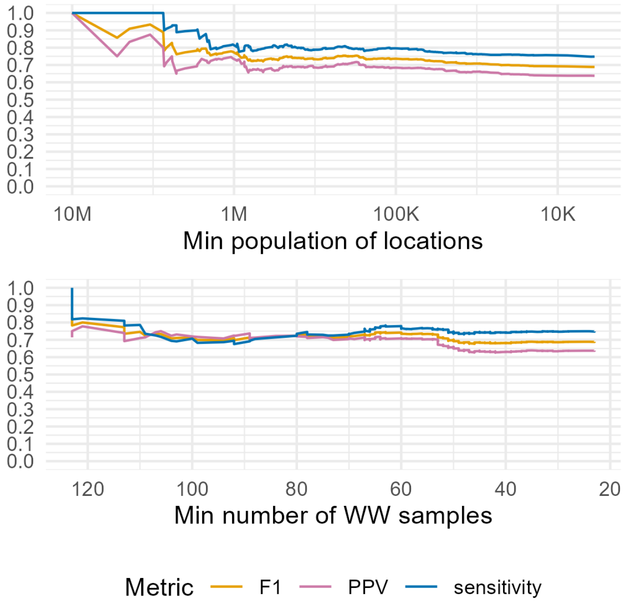


Figure S8: Rolling **outbreak detection** performance by location inclusion. Top: PPV, sensitivity, and F1 when locations are added in descending order of minimum population (x-axis, log scale; high population on the left). Bottom: same metrics when locations are added in descending order of minimum number of wastewater samples. Each point represents performance aggregated over all locations included up to that x-axis value.

[4] Jana S Huisman, Jérémie Scire, Daniel C Angst, Jinzhou Li, Richard A Neher, Marloes H Maathuis, Sebastian Bonhoeffer, and Tanja Stadler. Estimation and worldwide monitoring of the effective reproductive number of sars-cov-2. *Elife*, 11:e71345, 2022.

[5] Edward Goldstein, Jonathan Dushoff, Junling Ma, Joshua B Plotkin, David JD Earn, and Marc Lipsitch. Reconstructing influenza incidence by deconvolution of daily mortality time series. *Proceedings of the National Academy of Sciences*, 106(51):21825–21829, 2009.

[6] Anne Cori, Simon Cauchemez, Neil M Ferguson, Christophe Fraser, Elisabeth Dahlqwist, P Alex Demarsh, Thibaut Jombart, Zhian N Kamvar, Justin Lessler, Shikun Li, et al. Package 'epiestim'. *CRAN: Vienna Austria*, 13, 2020.
